# Development and evaluation of the SPACE-Postpartum multidomain symptom framework for predicting chronic pain after caesarean delivery: a prospective cohort study

**DOI:** 10.1101/2025.08.19.25333997

**Authors:** Sarah Ciechanowicz, Guillermina Michel, Nisha Panigrahy, Hannah Sukhdeo, Brendan Carvalho, Pervez Sultan

**Author notes:** Correspondence to: S Ciechanowicz. Data presented as Best Research Paper nomination, Society for Obstetric Anesthesiology and Perinatology, Portland, Oregon, USA, 1 May 2025.

## Abstract

**Background:** Chronic postsurgical pain after caesarean delivery impairs postpartum recovery and maternal quality of life. Existing risk models focus on demographic or procedural factors, limiting opportunities for early intervention. This study developed and prospectively evaluated a biopsychosocial predictive model, SPACE-Postpartum (Sleep, Pain, Affect, Cognition, Energy), which assesses early postpartum symptoms across five domains.

**Methods:** In this prospective cohort study, adults undergoing caesarean delivery at a tertiary centre completed validated patient-reported outcome measures at baseline, 2 weeks, and 3 months postpartum. Chronic pain was defined as pain at any site persisting >3 months. A five-item model with one early postpartum indicator from each SPACE domain was derived using logistic regression with ridge regularisation, supported by latent class and causal mediation analyses.

**Results:** Of 143 participants, 100 (70%) completed 3-month follow-up; 40% reported chronic pain. The SPACE ridge model demonstrated good discrimination (AUC 0.76, 95% CI 0.67–0.85) with internal validation and acceptable calibration. Higher acute pain intensity, pain interference, and sleep disturbance, with a trend for reduced perceived control, predicted chronic pain. Latent class analysis identified an early high-burden SPACE profile (52%) associated with greater pain interference at 3 months. Mediation analysis indicated acute pain exerted a direct effect, while sleep disturbance acted as an independent prognostic marker. Exploratory sensitivity analyses suggested potential incremental value of quadratic models (AUC 0.83–0.87), although these risked overfitting in this small dataset.

**Conclusions:** Chronic pain after caesarean delivery is common and linked to potentially modifiable early symptoms, particularly sleep disturbance and pain-related interference. Across predictive, phenotypic, and causal analyses, pain and sleep symptoms consistently demonstrated prognostic value. This single-centre proof-of-concept study provides early internal validation of the SPACE-Postpartum model. Multicentre external validation is warranted to confirm generalisability and support development of symptom-informed decision tools.

## Introduction

Persistent pain and poor recovery after childbirth represent major but under-recognised contributors to long-term morbidity. Chronic postsurgical pain (CPSP) following caesarean delivery (CD) affects an estimated 15-17% of individuals and contributes to reduced maternal wellbeing, impaired functioning, and diminished quality of life(1,2). Despite its prevalence, few strategies exist to identify those at greatest risk during the early postpartum period. In a recent meta-analysis, high-impact chronic pain, defined as pain that interferes with daily functioning, was found to affect up to 10% of patients after CD, underscoring the urgency of this issue(2).

The transition from acute to chronic pain is increasingly recognised as a multifactorial process involving biological, psychological, and behavioural influences(3–6). A 2019 review by Sun and Pan (7) identified diverse predictors of CPSP after CD, including preoperative factors (such as existing pain, anxiety, depression, psychological distress, and socioeconomic disadvantage), intraoperative variables (general anaesthesia, surgical complexity, postpartum haemorrhage), and postoperative features (severe acute pain, surgical complications, and depressive symptoms).

While these factors are well documented in the postpartum population, they are rarely incorporated into structured, theory-driven risk assessment models. Existing tools tend to focus on static demographic or procedural variables, with limited use of potentially modifiable, patient-reported symptoms(8). Only one CD-specific model to date includes early depressive and pain symptoms(9), but no existing framework integrates the full range of centrally mediated symptoms known to influence pain chronification.(10) This underscores the need for a comprehensive, mechanistically informed approach.

There is a need to move beyond siloed outcomes such as pain intensity or depression scores, toward the identification of actionable morbidity signatures - symptom constellations that are both predictive of poor outcomes and amenable to early intervention. In the postpartum context, where overlapping burdens of sleep disruption, mood disturbance, cognitive strain, and functional fatigue converge, such signatures could offer a scalable approach to anticipatory care. Recent advances in perioperative research underscore the potential of multidomain, symptom-based frameworks to capture recovery risk more holistically. The Stanford Obstetric Recovery Checklist (STORK) has recently been developed and validated as a structured tool for assessing postpartum recovery domains(11). The current study extends this line of enquiry by evaluating a multidomain framework for predicting chronic pain and impaired recovery after CD.

A recent narrative review highlighted that while the incidence of CPSP after CD is generally lower than after comparable gynaecological procedures, its true prevalence remains uncertain due to definitional challenges and methodological variability. The authors emphasised that ICD-11 criteria may not adequately capture the unique postpartum context, and that psychosocial and sleep-related factors are rarely assessed in existing studies(12). Furthermore, preventive strategies focused solely on surgical or analgesic techniques have yielded disappointing results, underscoring the multifactorial nature of CPSP. These limitations point to the need for multidimensional, postpartum-specific models of recovery that move beyond traditional biomedical predictors.

In recognition of the long-term impact of early life adversity on stress physiology and pain processing, adverse childhood experiences (ACEs) were included in this study as a preoperative psychosocial factor. Prior research has linked ACEs to heightened central sensitisation, dysregulation of the hypothalamic-pituitary-adrenal axis, and increased risk of chronic pain in adulthood, but have yet to be evaluated in the postpartum context(13).

## Aims and Hypothesis

This prospective cohort study aimed to identify early symptom predictors of chronic postpartum pain after CD. We hypothesised that pain, sleep disturbance and cognitive-affective symptoms in the early postpartum period could predict pain severity and interference beyond three months.

We further hypothesised that individuals with high early symptom burden across these domains would form a latent risk profile associated with persistent pain and impaired recovery.

## Methods

### Study Design and Participants

This was a prospective observational cohort study conducted at a tertiary academic medical centre, approved by the institutional Clinical Research Ethics Board (IRB-71824). Adult individuals (aged ≥18 years) undergoing scheduled or unscheduled CD between 28 February 2024 and 30 September 2024 were recruited using convenience sampling.

Exclusion criteria were: (1) inability to comprehend English or Spanish; (2) lack of capacity to provide informed consent; (3) gestational age <37 weeks; and (4) declined participation. Participants who did not complete baseline assessment or were lost to follow-up were considered to have discontinued participation. Patients were included irrespective of pre-existing chronic pain to reflect real-world postpartum heterogeneity. Participants received modest compensation for their time: US$20 for the 2-week follow-up and US$30 for the 3-month follow-up.

### Data Collection and Measures

Baseline demographics, obstetric history, and validated patient-reported outcome measures (PROMs) were collected: Adverse Childhood Experiences Questionnaire (ACE-Q), Brief Pain Inventory Short Form (BPI-SF), Edinburgh Postnatal Depression Scale (EPDS), State Trait Anxiety Inventory (STAI), Pain Catastrophizing Scale (PCS), PROMIS Sleep Disturbance Short Form 8a, and three novel items: ‘pain interference with ability to feel in control’, hereafter referred to as ‘perceived control interference’ and pain interference with the ability to feed, or hold baby. Follow-up was at 2 weeks and 3 months postpartum via REDCap surveys, with up to three reminders by SMS/telephone.

Chronic postpartum pain was defined as self-reported pain at any anatomical site persisting beyond 3 months, per BPI-SF. Anatomical pain locations were categorised as scar/abdomen/pelvis, back, or other.

### Framework Development and Scoring Approaches

This study was designed to both develop and internally validate a multivariable prediction model within a single cohort. The target sample size was estimated at approximately 130 participants, based on the anticipated incidence of chronic postpartum pain and the number of candidate predictors, consistent with recommendations for sample size in prediction model development(14).

The framework was derived directly from symptom data in this cohort across five domains (Sleep, Pain, Affect, Cognition, Energy). Predictive models and symptom profiles generated from these data formed the first operational prototypes of SPACE-Postpartum, which were then internally validated within the same cohort.

The SPACE-Postpartum framework was operationalised in this cohort using five symptom domains measured by validated PROMs: Sleep (PROMIS Sleep Disturbance Short Form 8a, 2 weeks postpartum), Pain (BPI-SF average pain intensity, 2 weeks), Affect (EPDS, 2 weeks), Cognition (perceived control interference, 24-48 hours), and Energy (BPI-SF interference with general activity, 2 weeks). PCS and STAI were also collected for exploratory analyses but were not retained in the final scoring tools due to weaker associations with chronic pain in this cohort.

Three main predictive scoring approaches and one exploratory model were developed and evaluated using chronic postpartum pain (pain at any site >3-months) as the primary outcome:

1. **Binary Prototype (v0.8):** Each domain dichotomised at predefined clinical thresholds; summed score range 0-5.
2. **Weighted Score (v0.9):** Continuous domain scores entered into multivariable logistic regression with linear and quadratic terms; regression coefficients applied as domain weights.
3. **Regularised Score (v0.9r):** Same predictors as v0.9 but without quadratic terms; ridge (L2) penalisation with optimism correction.
4. **Exploratory Quadratic Ridge (v0.9qr):** Penalised version of v0.9 with quadratic terms retained; tested for potential nonlinear effects but interpreted cautiously due to overfitting risk.

Given the modest sample size, this analysis was framed as proof-of-concept. Penalised logistic regression with internal validation was used to minimise overfitting, and supplementary analyses were treated as hypothesis-generating. We fit ridge (L2-penalised) logistic regression using scikit-learn with the regularisation parameter fixed at C=1.0 (inverse of λ). This approach was chosen to maintain stability in a small dataset and avoid potential overfitting from hyperparameter optimisation. Model performance was evaluated in terms of discrimination (AUC with 95% CI), calibration (calibration plots, slope, and intercept), overall accuracy (Brier score), and clinical utility (decision curve analysis). To account for potential overfitting in this modest sample, we prioritised internal validation using bootstrap resampling (1000 iterations), which is recommended as the most efficient method for optimism correction in small datasets.

The optimism-corrected C-statistic was considered our primary performance estimate. For transparency, we also performed 10-fold cross-validation, and a complete-case sensitivity analysis to evaluate the impact of imputation.

### Statistical Analysis

Descriptive statistics compared CPSP vs no-CPSP groups using Mann–Whitney U tests, χ²/Fisher’s exact tests, and Spearman correlations. Univariate logistic regression screened candidate predictors, although core SPACE domains were retained regardless of univariate significance. Multivariable models were then constructed and evaluated according to the predefined scoring approaches. Missing data were present in two domains (∼23%) but absent in the remaining three. Given the small dataset, missing values were handled using median imputation within the modelling pipeline (imputation → scaling → penalised logistic regression). This approach ensured that all participants could be included in model development, while recognising that more sophisticated methods (e.g. multiple imputation) may be preferable in larger datasets. Analyses were performed using R (version 4.3.2; R Foundation for Statistical Computing, Vienna, Austria) and Python (version 3.11; scikit-learn, pandas, numpy, matplotlib libraries).

### Latent Class Analysis

Latent class analysis (LCA) identified early symptom profiles using acute pain intensity (inpatient), sleep disturbance (2 weeks), and cognitive-affective interference (inpatient). A two-class model was selected via BIC. Class membership was related to 3-month pain interference using linear regression.

### Mediation Analysis

Causal mediation modelling examined whether sleep disturbance mediated the relationship between acute pain and chronic pain severity, adjusted for age, parity, prior chronic pain, and EPDS score.

## Results

### Participant Characteristics and Follow-up

A total of 143 participants were enrolled between February and September 2024. Follow-up was achieved in 105 participants (73%) at 2 weeks and 100 (70%) at 3 months postpartum (Supplementary Figure 1). 54 underwent planned CD and 46 intrapartum CD. There was no significant difference in CD type between participants with and without chronic postpartum pain (Pearson’s χ2 = 2.82, p = 0.093). Three patients (3%) at 3-month follow-up had a documented chronic pain condition (systemic sclerosis, congenital spinal abnormality, or endometriosis). Participant characteristics are described in Supplementary Table 1.

### Postoperative Pain Trajectories

Pain intensity declined over time (Supplementary Figure 2). At 24-48 hours post-CD, 53% of participants reported moderate pain (NRS 4–6), 33% mild pain (NRS 1-3), 10% severe pain (NRS 7-10), and 4% reported no pain (Supplementary Figure 3). By 2 weeks, 36% were pain-free, 45% had mild pain, 18% moderate pain, and 1% severe pain. At 3 months postpartum, 50% reported no pain, 26% mild pain, 19% moderate pain, and 5% severe pain. Chronic postpartum pain (defined as any daily pain at 3 months on the BPI) was reported by 40 participants (40%). Among these, the worst daily pain was mild in 20%, moderate in 52.5%, and severe in 27.5% (Supplementary Figure 4). Chronic pain interference with multiple subdomains of health-related quality of life is visualised in Figure 2.

**Figure 1.**
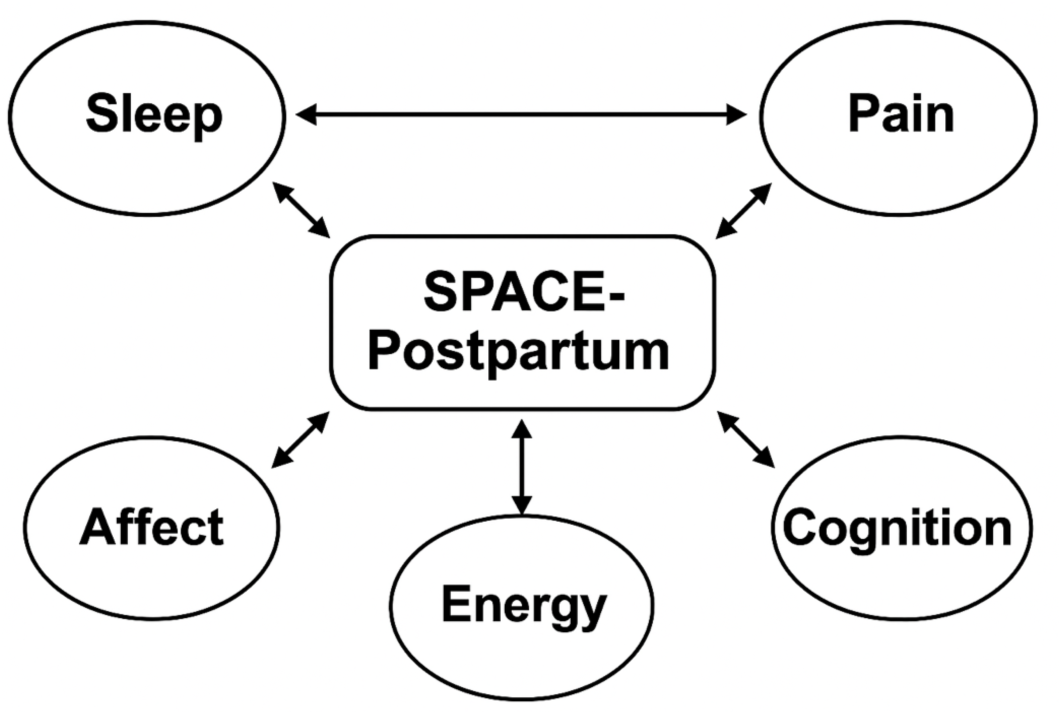
SPACE-Postpartum. A biopsychosocial framework for chronic postsurgical pain risk after caesarean delivery, comprising five interrelated symptom domains: Sleep, Pain, Affect, Cognition, and Energy. These domains are proposed as candidate mechanisms contributing to central pain vulnerability. Arrows indicate hypothesised bidirectional relationships.

**Figure 2.**
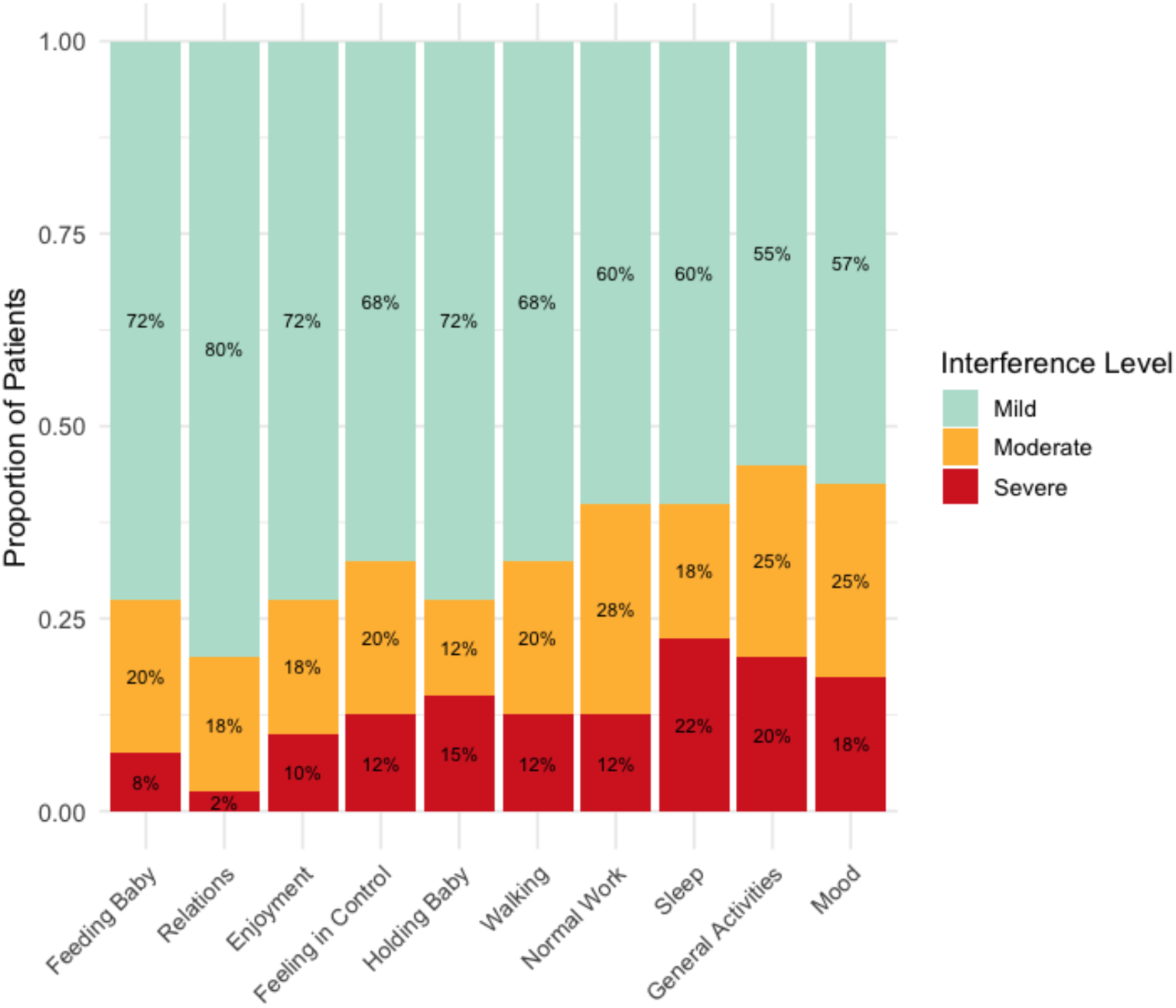
Pain interference across functional domains among participants reporting pain at 3 months postpartum. Bar chart shows the proportion of patients (n = 40) experiencing mild (0–3), moderate (4–6), and severe (7–10) interference in daily activities due to ongoing pain, based on self-reported ratings. Domains assessed included both infant-related activities (e.g., feeding, holding) and general functioning (e.g., walking, sleep, mood). The majority reported mild interference across domains, but 20-45% experienced moderate to severe interference in tasks such as walking, sleep, and general activity. Pain interference with feeding the baby and emotional well-being (e.g., mood, enjoyment) was also notable. This highlights the persistent functional impact of chronic postsurgical pain after caesarean delivery.

### Pain Distribution and Multisite Pain

At 3 months, the most common site of pain was the back (36%), followed by non-surgical regions (33%) and the scar/abdomen/pelvis (22%) (Supplementary Figure 5). Participants reporting chronic pain had a greater number of pain sites (median = 2, IQR 2–4) compared to those not reporting chronic pain (median = 1, IQR 1-1; Wilcoxon W = 278.5, p < 0.001). Multisite pain (≥2 sites) was reported by 95.7% of participants reporting pain sites at 3 months (Supplementary Table 2). While early multisite pain was not a significant predictor of chronic pain risk, the number of acute pain sites was significantly associated with the number of chronic pain sites reported at 3 months (Spearman’s rho = 0.20, p = 0.014), (Supplementary Figure 6).

In exploratory analyses, specific early symptom domains were associated with distinct chronic pain phenotypes. Greater inpatient interference with sleep (OR 1.35, 95% CI: 1.06–1.82, p = 0.024) and control (OR 1.51, 95% CI: 1.17–2.11, p = 0.006) were significantly associated with chronic scar or abdominal pain at 3 months (n = 22). Additionally, higher EPDS scores at 3 months were associated with scar-specific CPSP (scar, abdomen, or pelvis) (OR 1.11, p = 0.021), highlighting the continued impact of affective burden on pain persistence and vice versa.

### Psychological and Sleep-Related Correlates

Chronic postpartum pain at 3 months was associated with higher 24-hour postpartum state anxiety (STAI: ρ = 0.20, p = 0.020), depressive symptoms (EPDS: ρ = 0.18, p = 0.035), pain catastrophizing (PCS), and 2-week sleep disturbance (ρ = 0.27, p = 0.007). PCS was highly correlated with anxiety (ρ = 0.62, p < 0.001) and depressive symptoms (ρ = 0.54, p < 0.001).

Higher sleep disturbance at 2 weeks was associated with greater pain intensity and functional interference during the inpatient stay, including worst pain (β = 1.31, p = 0.003), current pain (β = 0.99, p = 0.026), general activity (β = 0.84, p = 0.007), and interpersonal relationships (β = 0.94, p = 0.023). At 3 months, 2-week sleep disturbance remained associated with interference in mood, work, relationships, sleep, and enjoyment of life (all p < 0.01). Sleep disturbance was associated with Edinburgh Postnatal Depression Scale (EPDS) scores and State-Trait Anxiety Inventory (STAI) scores at 2 weeks and 3 months (Supplementary Figure 7).

Higher EPDS scores were modestly associated with inpatient severe pain (β = 0.12, p = 0.038), worst (β = 0.12, p = 0.023) and average pain (β = 0.11, p = 0.006) at 2 weeks, but not with 3-month pain intensity. However, depressive symptoms were strongly associated with pain interference across timepoints, with the greatest effects at 2 weeks (e.g. enjoyment: β = 0.337, p < 0.001, R2 = 0.46); (Supplementary Table 3).

PCS was associated with greater inpatient worst pain (β = 0.076, p = 0.003) and functional interference. At 2 weeks, PCS predicted higher EPDS scores (β = 0.18, p < 0.001, R2 = 0.111) and sleep disturbance (β = 0.23, p = 0.022, R2 = 0.035). Associations with pain outcomes at 3 months were weaker and not statistically significant. Inpatient PCS were associated with anxiety, depression, and sleep disturbance outcomes (Supplementary Table 4).

State anxiety was not significantly associated with pain intensity or interference at any timepoint (all p > 0.05), although a weak trend was noted for interference with enjoyment of life at 2 weeks (β = –0.131, p = 0.054).

### Perceived Control

Greater pain-related interference with perceived control during the inpatient period was associated with greater pain severity at all timepoints. At 3 months, it was significantly associated with worst pain (β = 0.19, p = 0.027), average pain (β = 0.15, p = 0.028), and current pain (β = 0.24, p < 0.001).

### Adverse Childhood Experiences (ACEs)

Total ACEs score was not associated with pain intensity at any timepoint (all p > 0.25). However, ACEs were associated with greater inpatient interference with sleep (β = 0.594, p = 0.011, R2 = 0.064) and greater sleep disturbance at 2 weeks (β = 2.12, p = 0.005, R2 = 0.076), but not at 3 months (p = 0.469); (Supplementary Figure 8).

### Predictive Modelling

A multivariable logistic regression model using five early postpartum symptom domains from the SPACE-Postpartum framework - sleep disturbance, average pain intensity, pain interference with general activities (energy/fatigue), perceived control interference, and depressive symptoms - predicted chronic postpartum pain at 3 months with moderate/good discrimination.

A ridge-regularised model without quadratic terms (v0.9r) retained all five SPACE domains and was selected as the primary prototype for further validation (Supplementary Tables 5-6). Of the five predictors, three had complete data, while pain and energy both had 23% missingness; median imputation was applied (n = 23 of 100). The imputed SPACE ridge model demonstrated moderate/good discrimination, with an apparent AUC of 0.759 (95% CI 0.670–0.850, DeLong) (Fig.3). Apparent calibration was close to ideal (intercept = 0.037, slope = 1.095), and the Brier score was 0.204. Following internal validation with 1000 bootstrap resamples, the optimism-corrected AUC was 0.755 (95% CI 0.645–0.753). The optimism-corrected Brier score was 0.228 (95% CI 0.205–0.232). Calibration remained acceptable, with a corrected slope of 0.812 (95% CI 0.509–1.364) and an intercept of –0.056 (95% CI –0.427 to 0.499). For sensitivity analysis, notably, 10-fold cross-validation yielded a somewhat lower mean AUC of 0.68 (SD 0.17), likely due to the high variability in this small sample; we relied on the bootstrap optimism-corrected estimate as a more reliable indicator of performance. A complete-case analysis (n = 77) produced an AUC of 0.76. Given the small sample, we used median imputation for simplicity; this likely shrinks variance and could bias associations toward null. However, the concordant complete-case analysis supports that our findings were not an artefact of the imputation method.

**Figure 3.**
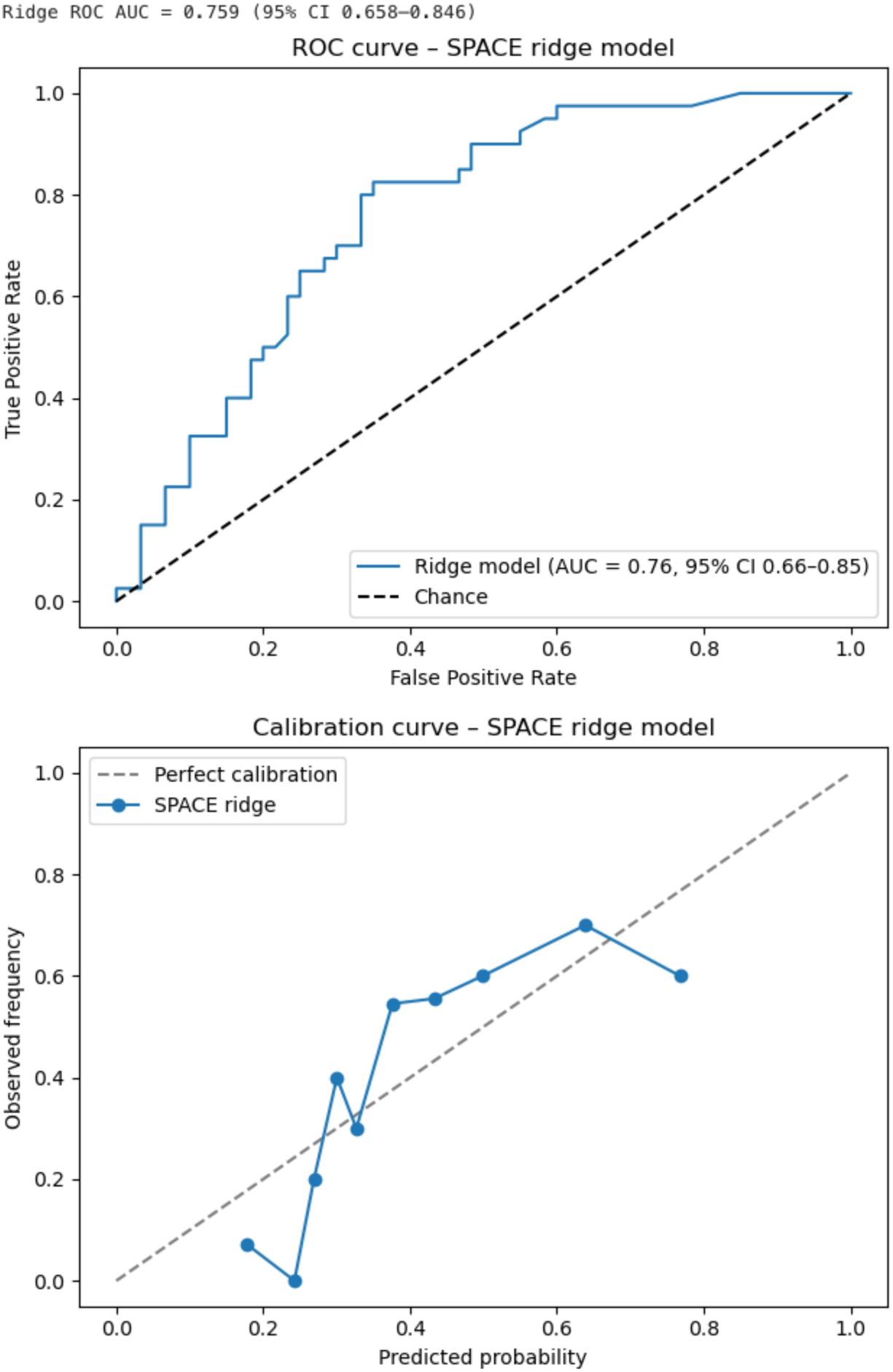
Receiver operating characteristic (ROC) curve for imputed ridge SPACE score (v0.9r) predicting chronic postsurgical pain at 3 months. Demonstrated good discrimination for chronic pain (AUC = 0.76), based on symptom domain scores at baseline and two weeks postpartum. True positive rate is plotted against false positive rate for model-predicted risk across thresholds. The model was derived from five symptom domains (Sleep, Pain, Affect, Cognition, and Energy).

An unregularised quadratic model (v0.9) achieved higher apparent discrimination (AUC = 0.83; 95% CI 0.73–0.92) with acceptable calibration (Hosmer–Lemeshow p = 0.07). A quadratic-augmented ridge model yielded an AUC of 0.87 (Supplementary Fig.9), although this likely reflects model complexity in a modest dataset and should be interpreted with caution pending external validation. Replacing depressive symptoms with BMI reduced discrimination (AUC = 0.77), underscoring the prognostic importance of affective symptoms.

Together, these findings support the internal validity of the SPACE ridge model, with stable discrimination (AUC 0.76) despite the modest sample size.

### Prototype Scoring and Feasibility

Four approaches were compared: v0.8 (binary), v0.9 (weighted quadratic), v0.9r (ridge-regularised linear), and v0.9qr (ridge-regularised quadratic). The exploratory v0.9qr achieved the highest apparent AUC (0.87) but was likely overfitted (Supplementary Figure 9). For clinical translation, v0.9r was prioritised for its generalisability and interpretability, while v0.8 offers a pragmatic low-burden screening option (Supplementary Table 6). To explore clinical utility, we performed a supplementary decision curve analysis (DCA). The SPACE-Postpartum ridge model demonstrated net benefit across threshold probabilities of ∼0.20-0.60, supporting potential value for risk stratification in early postpartum care (Supplementary Fig. 10).

### Latent Class Analysis

We focused on three LCA indicators that captured pain, sleep, and cognitive-affective control; adding more domain variables was not feasible due to sample size and model stability. A two-class latent model identified a low-burden SPACE class (48%) and a high-burden SPACE class (52%), based on early sleep disturbance, pain, and perceived control. Participants in the high-burden class had significantly higher 3-month least pain (mean 1.71 vs 0.90, β = 0.81, p = 0.043) and average pain interference (mean 1.74 vs 0.86, β = 0.88, p = 0.024); (Figure 4).

**Figure 4.**
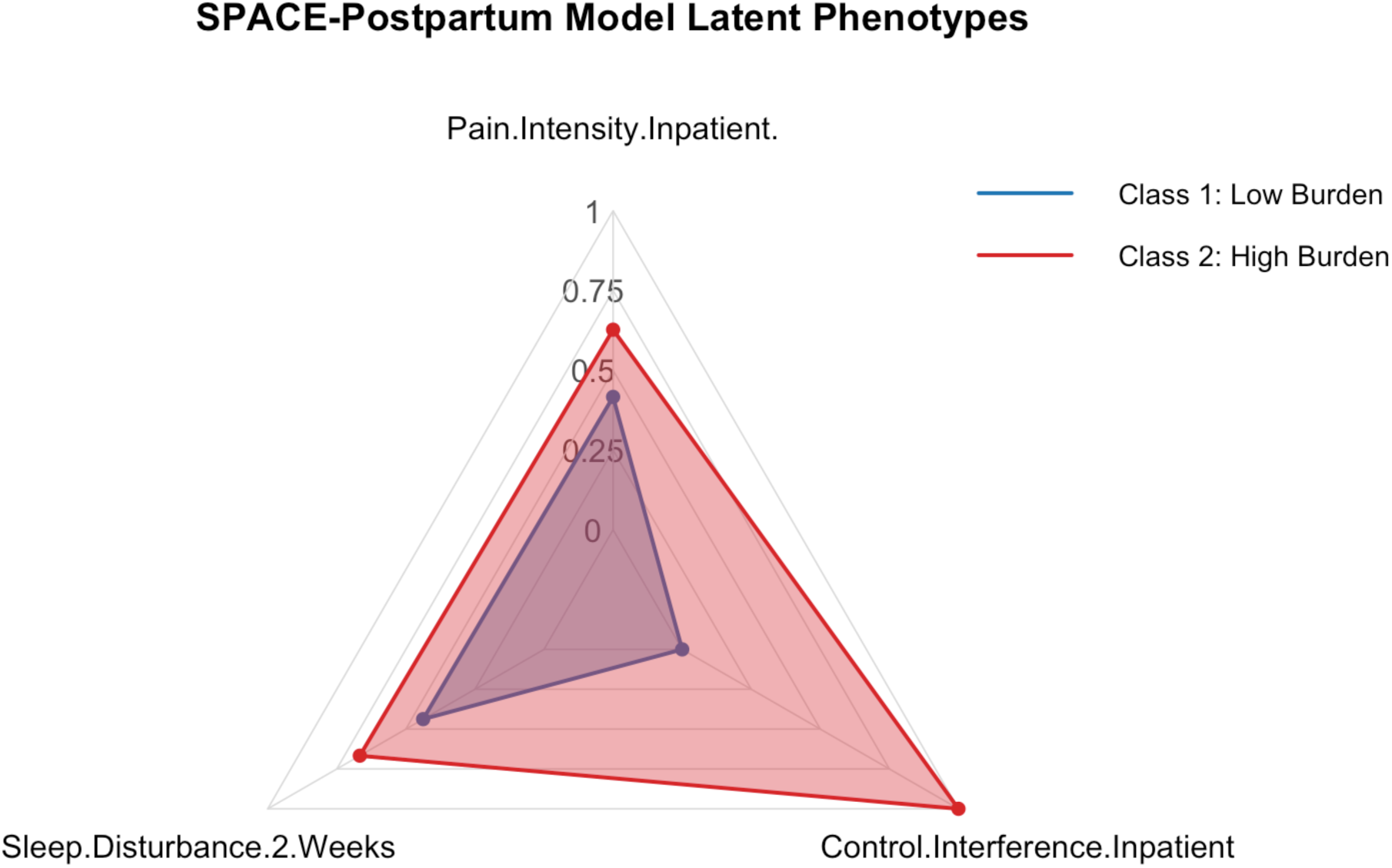
Latent symptom profiles derived from the SPACE framework and their association with 3-month pain outcomes. Two latent classes were identified using a three-domain SPACE model incorporating early postpartum symptoms: sleep disturbance at two weeks, pain intensity within 24–48 hours, and perceived cognitive-affective interference (perceived control) within 24–48 hours. Class 2 (red) represents a high-burden phenotype with elevated conditional probabilities across all three domains; Class 1 (blue) represents a low-burden phenotype. Values are scaled from 0 to 1 and reflect class-specific probabilities of symptom presence. The model was based on 99 complete cases and significantly predicted 3-month pain outcomes: Class 2 membership was associated with higher least pain intensity (β = 0.81, 95% CI: 0.02–1.60, p = 0.043) and greater average pain interference (mean 1.74 vs 0.86; β = 0.88, 95% CI: 0.12–1.64, p = 0.024), explaining 5.2% of variance in pain-related interference (adjusted R² = 0.042). These findings suggest that early clustering of symptoms across SPACE domains may help identify individuals at risk of persistent pain-related functional impairment after caesarean delivery. This high-burden latent class is hereafter referred to as the SPC (Sleep–Pain–Cognition) profile.

### Mediation Analysis

Acute pain severity was significantly associated with chronic pain at 3 months (Average Direct Effect = 0.32, 95% CI 0.09–0.55, p = 0.0068). Two-week sleep disturbance partially attenuated this relationship, but the indirect (mediated) effect was not statistically significant (Average Causal Mediation Effect = 0.025, 95% CI –0.049 to 0.11, p = 0.51). A composite SPACE burden score yielded similar findings. These results suggest that early sleep disturbance contributes to pain trajectories but does not statistically mediate the acute-to-chronic transition in this cohort (Supplementary Figure 11; Supplementary Table 7).

## Discussion

### Principal Findings

This study was designed as an exploratory, proof-of-concept analysis to test the feasibility of a multidimensional biopsychosocial prediction framework (SPACE) for chronic postsurgical pain after CD. Given the modest sample size and event rate, our modelling strategy prioritised parsimony and internal validity: we applied penalised (ridge) logistic regression to reduce overfitting, assessed performance with bootstrap cross-validation, and reported discrimination, calibration, and Brier score as recommended. While supplementary analyses (e.g. latent class symptom profiling, decision curve analysis) were undertaken to illustrate potential clinical and mechanistic insights, these should be interpreted as hypothesis-generating. External validation in larger, multicentre cohorts will be required before the SPACE model can be considered for clinical application.

We found that 40% of participants reported persistent pain three months after CD, underscoring the significant burden of chronic pain in postpartum populations. Key early predictors of chronic pain included higher acute pain intensity and interference, sleep disturbance, and indications of cognitive-affective interference - factors collectively captured within the symptom-based SPACE-Postpartum framework(15), which integrates Sleep, Pain, Affect, Cognition, and Energy (fatigue) domains (Fig. 1). The framework, derived and applied both conceptually and analytically, demonstrated that early symptom burden predicted pain at three months, supporting the feasibility of symptom-based early risk stratification. Notably, a high symptom burden latent phenotype was identified in just over half the cohort, associated with worse pain outcomes, highlighting clinical heterogeneity.

We developed and internally validated a novel biopsychosocial framework. SPACE domains are known to contribute to pain central sensitisation, be prevalent in postpartum populations, and have the potential to serve as modifiable targets for early intervention. While similar symptom clusters have been described in other chronic pain populations (16–18), the SPACE-Postpartum model represents the first structured, mechanistically informed application of such a model to postpartum surgical recovery. The framework is supported by neurobiological evidence linking mood, sleep, and cognitive disruption to altered activity in the prefrontal cortex, amygdala, and anterior cingulate cortex(19). Fatigue and low energy are associated with hypothalamic-pituitary-adrenal axis dysfunction and systemic inflammation. Together, these mechanisms contribute to central sensitisation, impaired descending inhibition, and neuroimmune activation, key processes in the chronification of pain(20).

### Interpretation and Conceptual Context

These results reinforce the biopsychosocial model of pain chronification, emphasising the pivotal role of central nervous system symptom burden, particularly early disruptions in sleep, mood, and cognitive-affective functioning, in the transition from acute to chronic pain. The SPACE-Postpartum framework advances existing models by providing a structured, patient-centred approach to capture modifiable early symptoms.

A novel finding was the value of perceived interference with control, a cognitive-affective disruption measure developed specifically for this study. This predictor outperformed traditional instruments such as the Pain Catastrophizing Scale (PCS), which, while associated with acute distress, lost predictive strength over time, suggesting it reflects more transient psychological states. It was chosen because ObsQoR item ‘I feel in control’ had an apparent association with 6-week EPDS scores in a prior study(22). Its strong signal here suggests it taps into an important domain (sense of control) that merits further validation in future studies.

Sleep disturbance measured at two weeks postpartum consistently predicted CPSP in both multivariable and latent class analyses, aligning with mechanistic evidence linking poor sleep to neuroinflammation, impaired descending pain modulation, and affective dysregulation, key contributors to nociplastic pain. Although mediation analyses did not identify significant indirect pathways via sleep or affective symptoms, the direct association between acute pain and CPSP highlights the urgency of optimised early analgesia. Consistent with Hayes (2013), nonsignificant mediation results in modestly powered studies should be interpreted cautiously and do not preclude underlying mechanistic processes(21).

Importantly, symptom-based tools like SPACE may mitigate bias and resource limitations inherent in traditional risk models that rely on demographic or procedural factors, which can encode structural inequities. By focusing on patient-experienced symptoms, the framework offers a more equitable perspective on postpartum recovery.

Unlike recent EHR-based models such as Liu et al. (2025)(8), which mainly depend on static demographic and procedural variables and often act as “black-box” predictors, the SPACE-Postpartum framework employs prospectively collected, modifiable symptom domains that can be monitored and addressed in real time.

### Strengths and Limitations

Key strengths of this study include its prospective design, comprehensive symptom profiling across multiple domains, and the combined use of predictive modelling and latent class analysis to interrogate the SPACE framework. The inclusion of novel cognitive-affective measures and repeated assessments enhances mechanistic understanding. The principal limitation of this study is its modest sample size, which inevitably constrains model precision and precludes external validation within this dataset. However, this should be viewed in the context of the study’s proof-of-concept design: the primary aim was to demonstrate feasibility of the SPACE-Postpartum framework and to explore its mechanistic coherence across sleep, pain, affect, cognition, and energy domains. Importantly, our modelling strategy incorporated ridge penalisation, imputation, and bootstrap optimism-correction, methodological safeguards that are often absent from larger published CPSP models, which frequently rely on unregularised regression and limited calibration assessment. Thus, while the present findings require confirmation in larger, multicentre cohorts, they provide promising preliminary evidence that symptom-based, penalised modelling is both feasible and promising for postpartum CPSP risk stratification. The latent class analysis findings also require replication in larger, external cohorts. Additionally, the Energy domain was operationalised via pain interference with activities rather than a dedicated fatigue measure, limiting the framework’s completeness.

Although Affect (measured via the EPDS) did not emerge as a strong predictor of chronic pain, it was retained within the SPACE framework due to its theoretical importance and consistent associations with poor long-term recovery. Large-scale evidence from the UK Biobank reinforces this decision: among 70,630 general adults without baseline pain, mood disturbance was one of three core central nervous system-driven symptoms, alongside sleep disturbance and cognitive disruption, associated with the later development of chronic primary pain (16).

Qualitative interviews of participants experiencing chronic pain from our own study revealed lived experiences closely aligned with the SPACE domains, including disrupted sleep, affective distress, diminished control, and fatigue, adding further interpretive and patient-centred validity. This triangulation across prospective cohort data, qualitative interviews, and population-level risk modelling highlights Affect as a mechanistically meaningful and clinically actionable domain, even where short-term predictive strength may vary by timepoint or cohort.

The SPACE symptom cluster identified in the UK Biobank as a significant predictor of chronic pain onset in the general population suggests this signature may represent a common vulnerability phenotype rather than one confined to obstetric populations(16), but there could be important differences that require further study. To our knowledge, this study is the first to operationalise a SPACE early morbidity signature into a structured, regularised risk prediction tool for early postoperative use. Collectively, this triangulation of findings support the theoretical coherence, ecological validity, and translational potential of SPACE as a dynamic, symptom-based framework for identifying and addressing poor recovery trajectories.

Notably, SPACE is designed as a dynamic framework, recognising that different symptom domains may exert differential influence at distinct recovery phases. While sleep and pain intensity dominate early risk prediction, depressive symptoms may become increasingly salient later in the postpartum course, particularly in relation to fatigue, pain interference, bonding, and return to function. Future longitudinal modelling may clarify these temporal dynamics and support personalised interventions at appropriate recovery stages.

Unlike traditional CPSP definitions confined to the surgical site, our outcome captured any persistent pain, aligning with a holistic view of postpartum recovery. This broader definition likely explains the higher incidence (40%) observed. We chose a broad definition of chronic pain to reflect overall maternal morbidity; however, this could introduce heterogeneity in the outcome. Future studies might distinguish pain likely attributable to the surgical procedure from other sources of postpartum pain. Our observed prevalence of documented chronic pain (3%) was lower than that reported in other antenatal cohorts, where 10-28% of women had a formal chronic pain diagnosis(23). This likely reflects our reliance on recorded comorbidities rather than structured pain assessments, and therefore represents a conservative estimate.

We performed multiple exploratory analyses; these were not corrected for multiple comparisons, so some associations (especially marginal ones) should be interpreted with caution.

### Clinical Implications

The findings support the potential of early symptom-based screening to identify individuals at elevated risk of CPSP after CD. Modifiable symptoms such as sleep disturbance, perceived loss of control, mood, and pain interference are measurable in routine care and amenable to interventions. The regularised model’s good predictive accuracy (AUC = 0.759; optimism-corrected 0.755) underlines the feasibility of integrating symptom-based risk stratification into postpartum pathways.

Across surgeries, published CPSP models report AUCs spanning 0.74-0.90, with performance depending on predictors and validation strategy: externally validated models often fall around 0.74-0.82 (breast cancer model externally validated at 0.74; a mixed-surgery model that included 2-week postoperative pain/sensory features reported AUC = 0.82 with external confirmation)(24,25). Higher AUCs (0.88–0.90) are typically from internal-only nomograms or ML models in specific cohorts (total knee arthroplasty; postpartum retrospective XGBoost)(8,26), which may attenuate with external testing. In contrast to many prior tools that rely on static demographic/procedural factors and unpenalised modelling, the SPACE-Postpartum model integrates dynamic, patient-reported, modifiable symptoms collected early postpartum and applies ridge regularisation with optimism correction, achieving discrimination within the externally validated range while enhancing interpretability and modifiability (Supplementary Table 8).

A key strength of SPACE-Postpartum is its parsimony: five domains operationalised with a single item or PROM per domain, enabling efficient risk stratification without overwhelming patients or clinicians. This minimalist structure enhances feasibility for clinical integration while maintaining robust predictive accuracy. Compared with STORK, which provides a comprehensive assessment of postpartum recovery(11), SPACE-Postpartum offers complementary predictive value by focusing on early, modifiable symptoms. For SPACE-Postpartum v0.9r, a three-tier risk classification identified ∼25% as high risk (>0.60 predicted probability), consistent with thresholds used in comparable tools and supporting targeted early intervention.

Adverse childhood experiences (ACEs) were significantly associated with greater early postpartum sleep disturbance, suggesting a potential vulnerability pathway through which early-life stress may dysregulate stress-response and sleep systems, contributing to poorer postpartum recovery and increased risk of worse pain and mood outcomes.

### Future Research

External validation of the SPACE-Postpartum framework and triage prototype in larger, diverse cohorts, including secondary analyses of larger datasets and multi-centre studies is needed.

Mechanistic studies exploring biological correlates of symptom burden, particularly the role of sleep in pain vulnerability, could refine intervention targets. Comparative investigations of cognitive-affective constructs like perceived control versus catastrophizing may enhance framework optimisation. Integration of symptom-based models into electronic health records and digital health technologies in postpartum care pathways has the potential to personalise recovery trajectories and reduce disparities in pain-related outcomes.

## Conclusion

CPSP following CD is common and under-recognised, with significant impacts on maternal functioning and quality of life. This study provides prospective evidence that early postpartum symptom burden - especially sleep disturbance, pain burden, and cognitive-affective interference - predicts chronic pain development. The SPACE-Postpartum framework could offer a novel, patient-centred approach to risk stratification and early intervention. By centring on patient experience, it may promote equitable care and improve long-term maternal outcomes.

The current analysis represents an early, proof-of-concept internal validation of the SPACE-Postpartum model within a single cohort, and findings require confirmation in diverse external populations to ensure generalisability. Future work should focus on multi-centre external validation, refinement of predictive thresholds, and integration into symptom-informed decision support tools that could contribute to more proactive postpartum care. Unlike traditional tools that measure isolated outcomes, SPACE conceptualises recovery as a dynamic interplay of interdependent domains, each of which may shift in salience over time. This first operationalisation of a SPACE morbidity signature into a structured predictive model (SPACE-Postpartum v0.9r) demonstrates the feasibility of using validated and novel patient-reported measures. Embedding SPACE into a regularised, clinically usable tool could lay the foundation for targeted postpartum risk stratification, symptom-informed recovery, and potential future integration into AI-driven clinical decision support systems.

## Data Availability

All data produced in the present study are available upon reasonable request to the authors

## Acknowledgements

We thank the participants who generously shared their experiences for this study, and Nan Guo for statistical analysis support. AI software (ChatGPT, OpenAI, San Francisco, CA) was used to support language editing and streamline code formatting. All statistical analyses and interpretations were performed independently by the authors.

## Author Contributions

S.C. conceptualised the SPACE framework and led study design, data collection, analysis, and manuscript drafting. G.M. contributed to participant recruitment, data acquisition, data management and manuscript revision. N.P. and H.S. contributed to participant recruitment, data acquisition and manuscript revision. B.C. contributed to supervision, cohort study design, interpretation and manuscript revision. P.S. contributed to supervision, cohort study design, interpretation and manuscript revision.

## SUPPLEMENTARY FILES

**Supplementary Figure 1.**
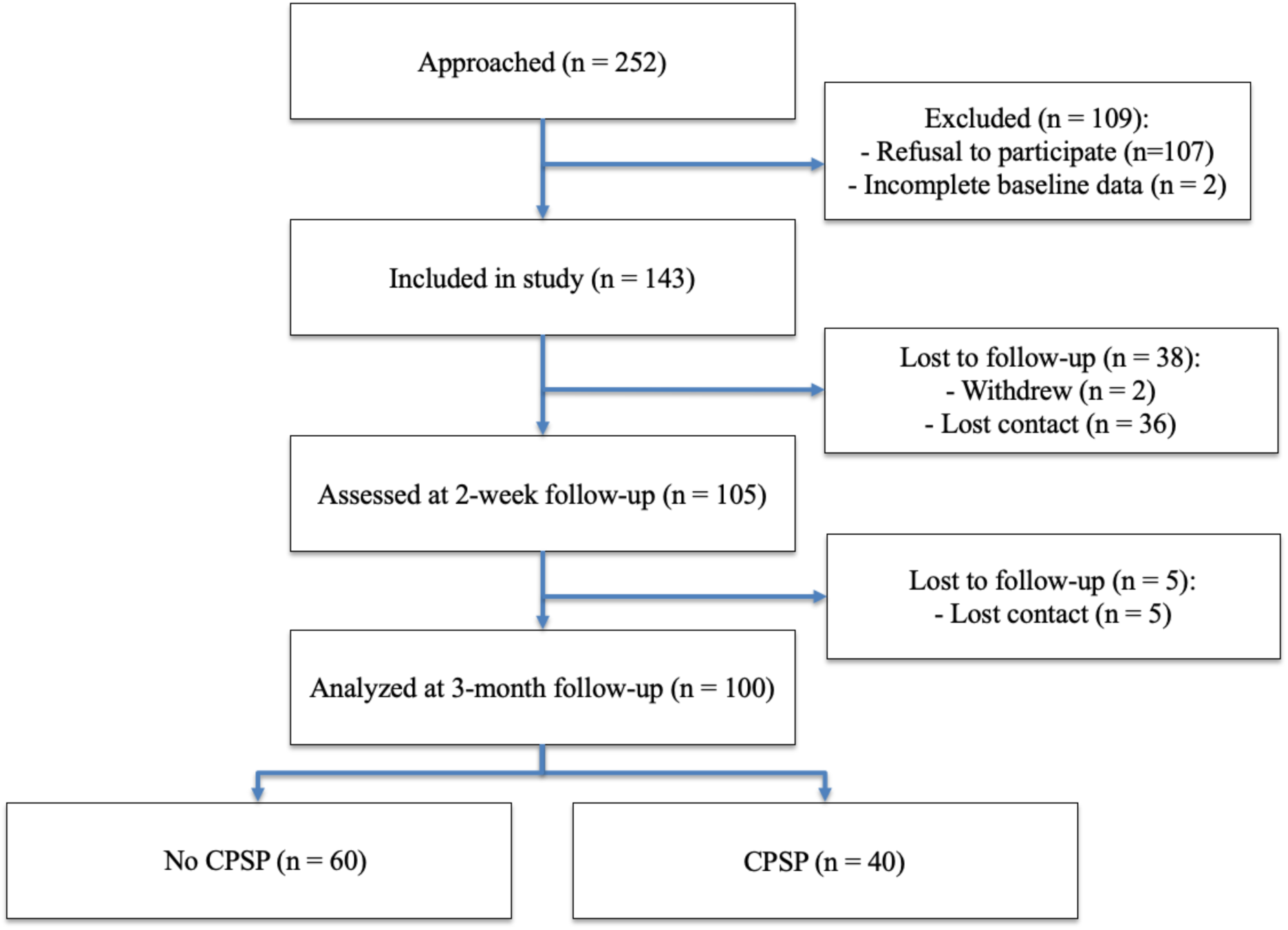
Study flow diagram (CONSORT). CPSP = chronic postsurgical pain (all sites). All postpartum individuals who underwent caesarean delivery were pre-screened using electronic healthcare records. The vast majority were eligible; only those not meeting inclusion criteria (e.g. age <18, language barrier, major comorbidity) were excluded at this stage. Only pre-eligible women were approached for participation. The absolute number screened in records was not separately logged.

**Supplementary Figure 2.**
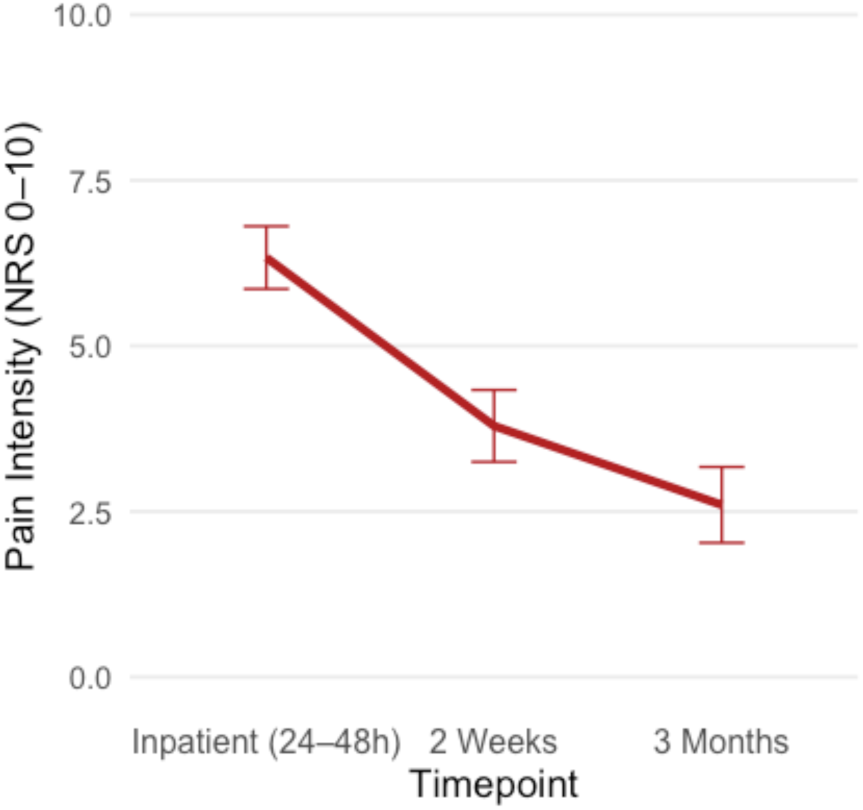
Mean pain intensity scores over the study duration.

**Supplementary Figure 3.**
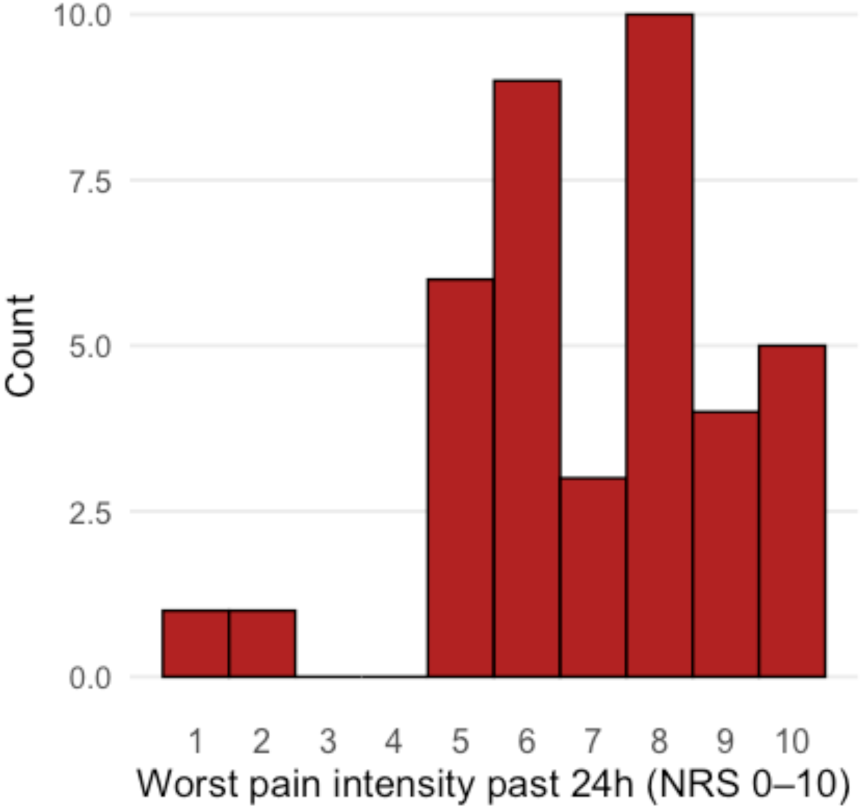
Distribution of worst pain intensity scores during the past 24 hours among participants reporting pain at 24 to 48 h postpartum. Pain intensity was measured on a 0–10 numerical rating scale.

**Supplementary Figure 4.**
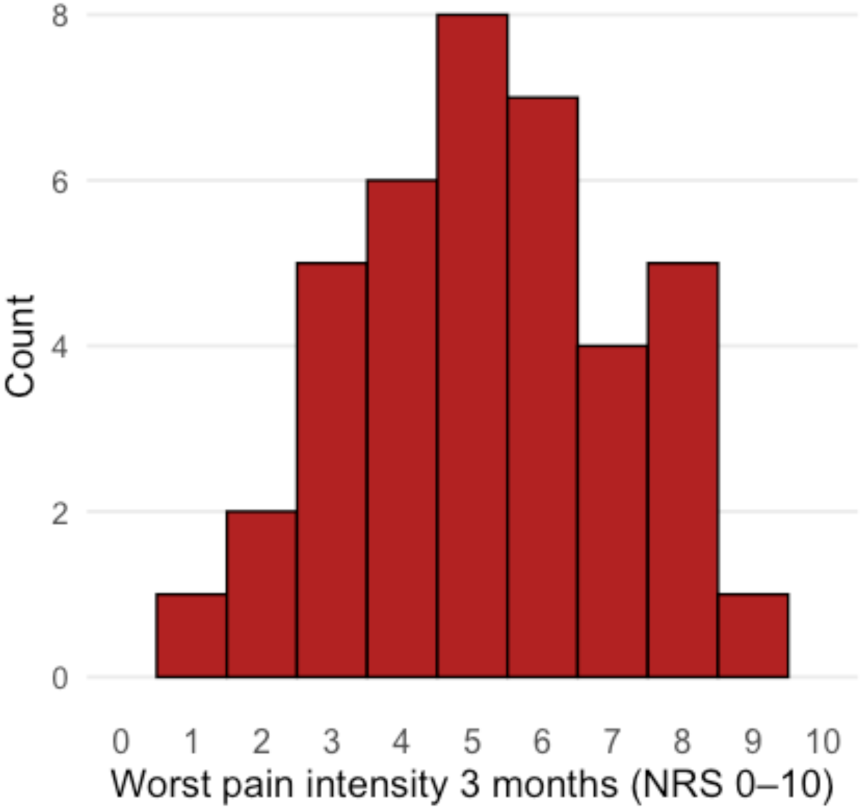
Distribution of worst pain intensity scores during the past 24 hours among participants reporting pain at 3 to 4 months postpartum. Pain intensity was measured on a 0–10 numerical rating scale.

**Supplementary Figure 5.**
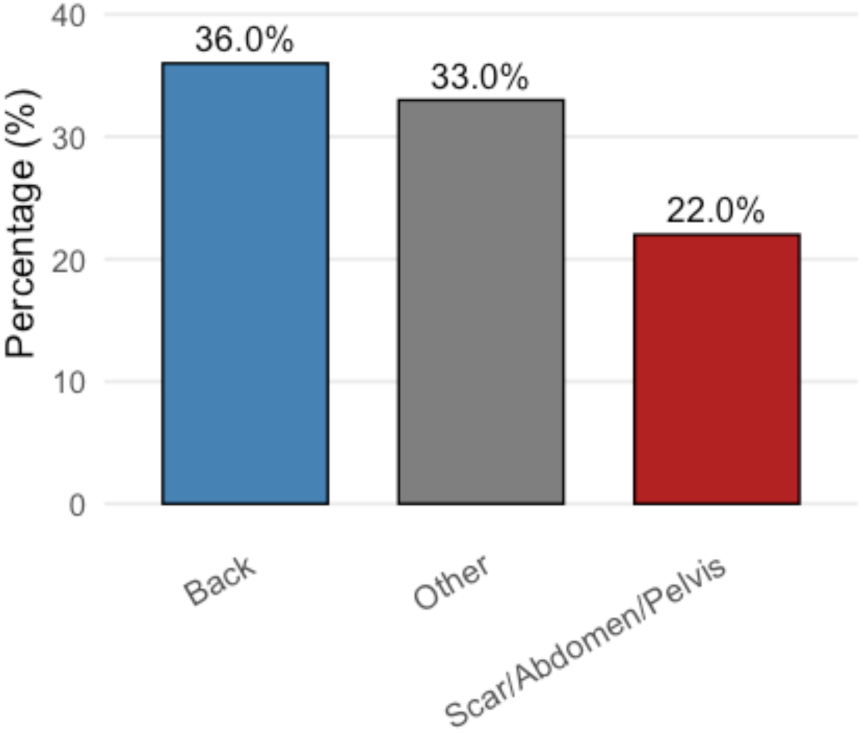
Percentage of participants reporting chronic pain by body region at 3-4 months postpartum. Chronic pain locations were categorised into Scar/Abdomen/Pelvis (surgical wound area), Back (thoracic and lumbar regions), and Other anatomical sites (head, neck, legs, arms, hands). Percentages represent the proportion of participants reporting pain in each region.

**Supplementary Figure 6.**
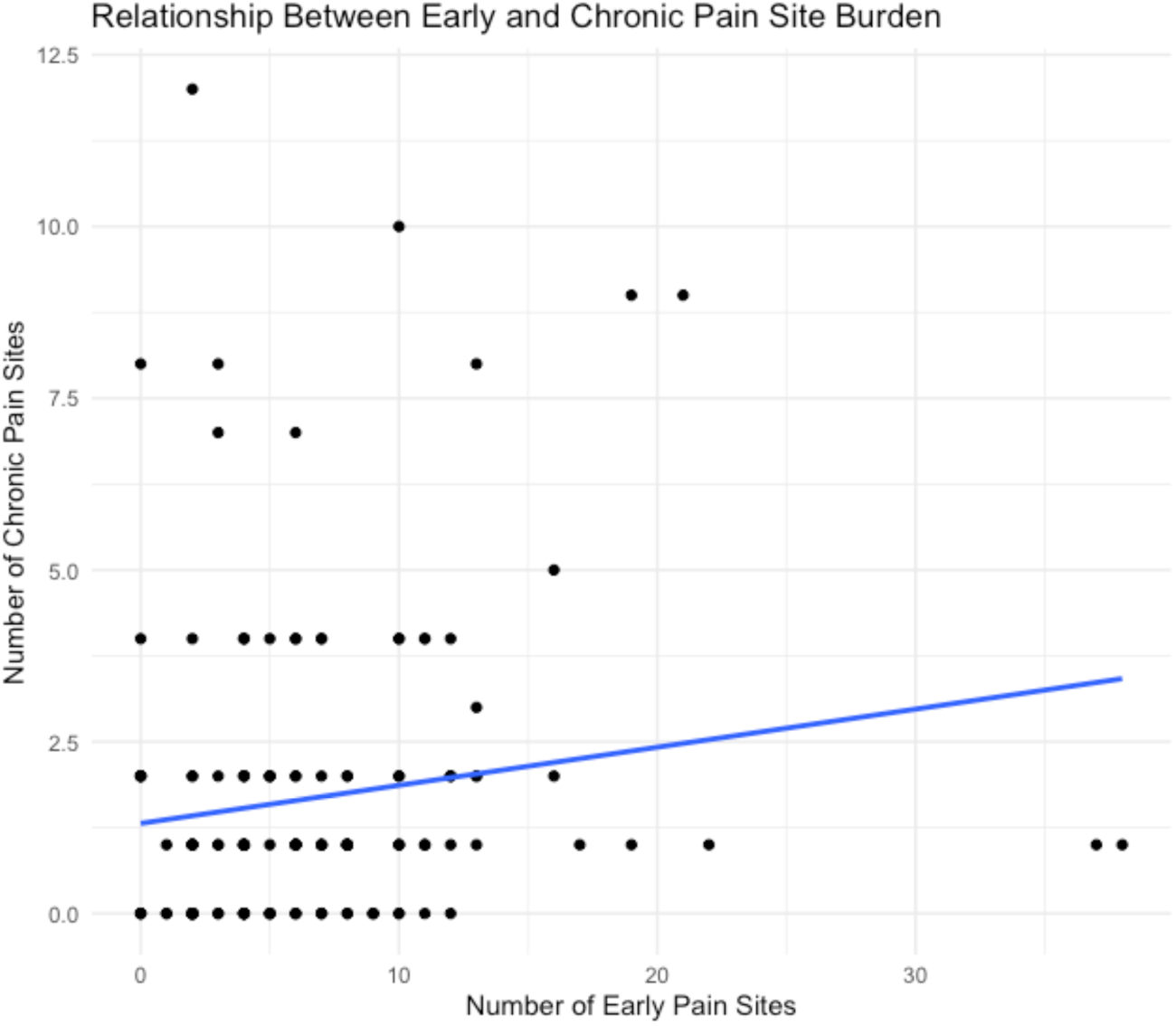
Relationship between the number of acute inpatient postpartum pain sites and the number of chronic pain sites at three months following caesarean delivery. Each dot represents an individual participant. A positive association was observed (Spearman’s rho = 0.20, p = 0.014).

**Supplementary Figure 7.**
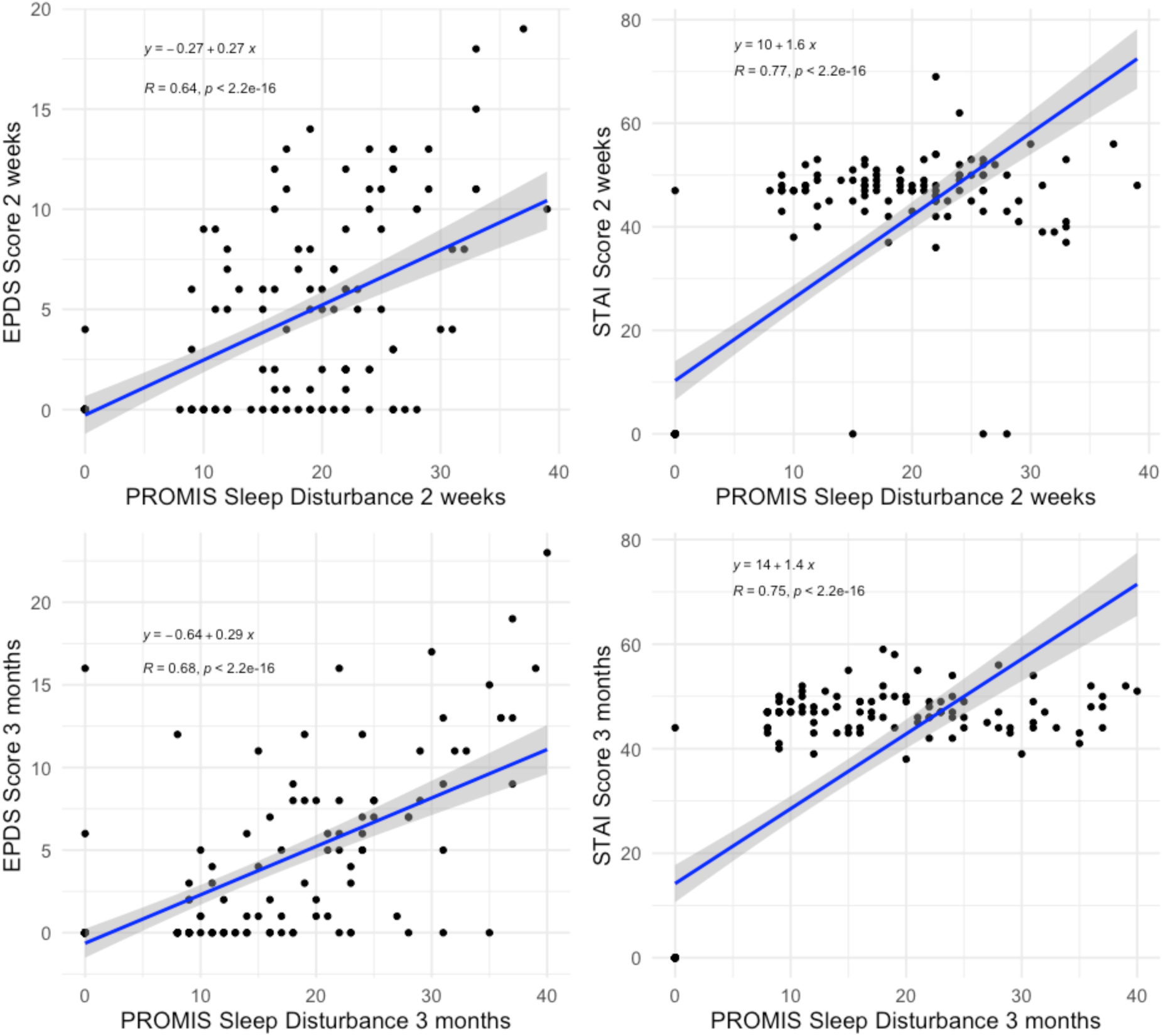
Association between PROMIS Sleep Disturbance scores and psychological symptom severity postpartum. Scatterplots illustrate linear relationships between sleep disturbance and Edinburgh Postnatal Depression Scale (EPDS) scores and State-Trait Anxiety Inventory (STAI) scores at 2 weeks (top row) and 3 months (bottom row) following caesarean delivery. Regression lines with 95% confidence intervals are shown. Equations, R² values, and Pearson correlation coefficients (r) with associated p-values are displayed on each panel.

**Supplementary Figure 8.**
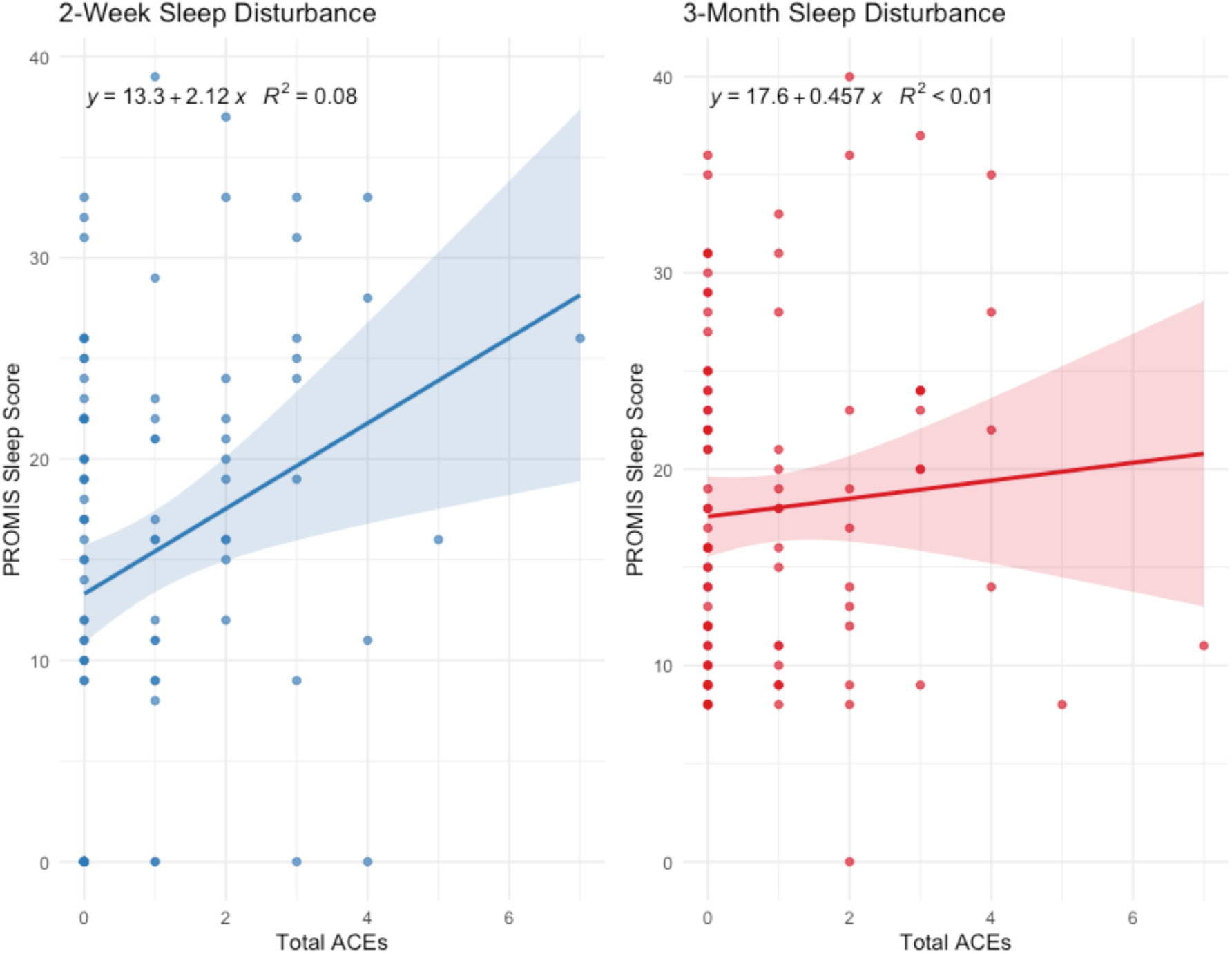
Association of Adverse Childhood Experiences (ACEs) score on sleep disturbance scores at 2 weeks and 3 months postpartum. P=0.005 at 2 weeks, indicating a significant positive relationship at this time-point. 3 month association was non-significant (P>0.05).

**Supplementary Figure 9.**
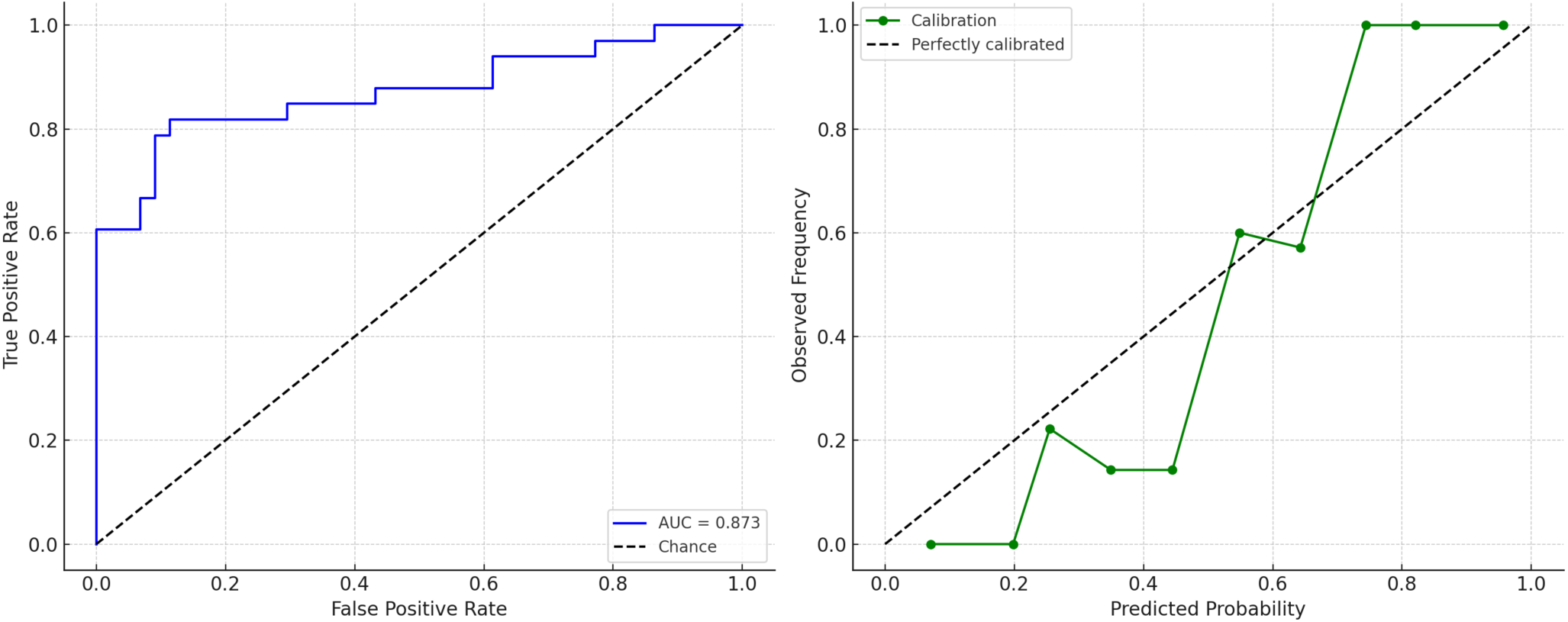
Discrimination and calibration of the weighted SPACE quadratic ridge model (v0.9rq) for predicting chronic postsurgical pain (CPSP) at 3 months after caesarean delivery. Left panel: Receiver operating characteristic (ROC) curve for the SPACE-Postpartum model (v0.9rq), developed using quadratic ridge regression to account for non-linear relationships and reduce overfitting. The model included five early symptom-based predictors and demonstrated high discrimination (AUC = 0.87). Right panel: Calibration curve comparing predicted probabilities with observed CPSP frequencies. While overall model fit appears strong, deviations from perfect calibration suggest potential overfitting, likely due to the sample size and model complexity. The dashed diagonal line indicates perfect calibration.

**Supplementary Figure 10.**
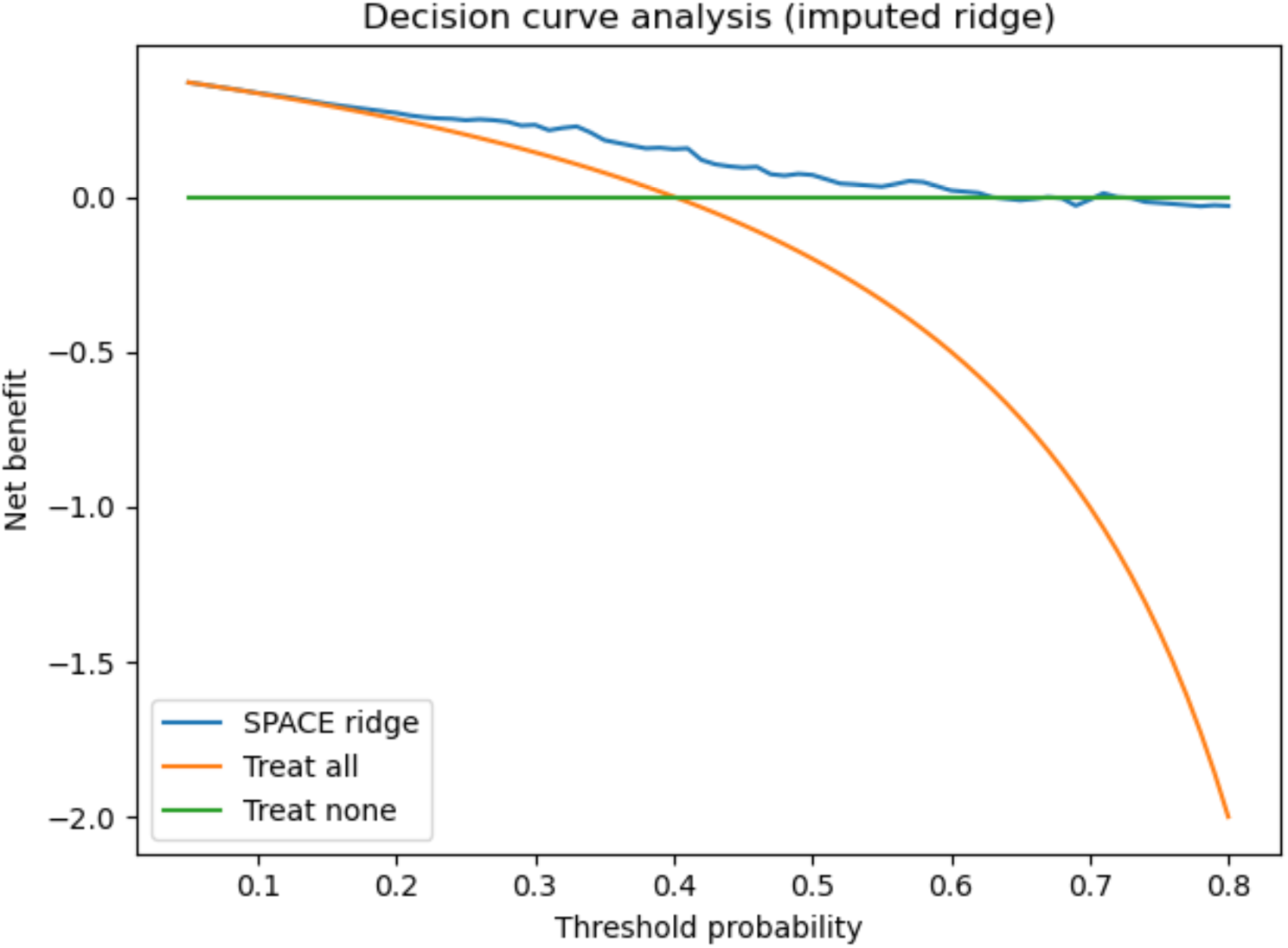
Decision curve analysis of the SPACE ridge model (v0.9r, imputed dataset) for predicting chronic postsurgical pain at 3 months postpartum. The SPACE ridge model (blue line) is compared with “treat all” (orange line) and “treat none” (green line) strategies across threshold probabilities from 0.05 to 0.80. The model provides a higher net benefit than either default strategy within clinically relevant decision thresholds (approximately 0.2-0.6).

**Supplementary Figure 11.**
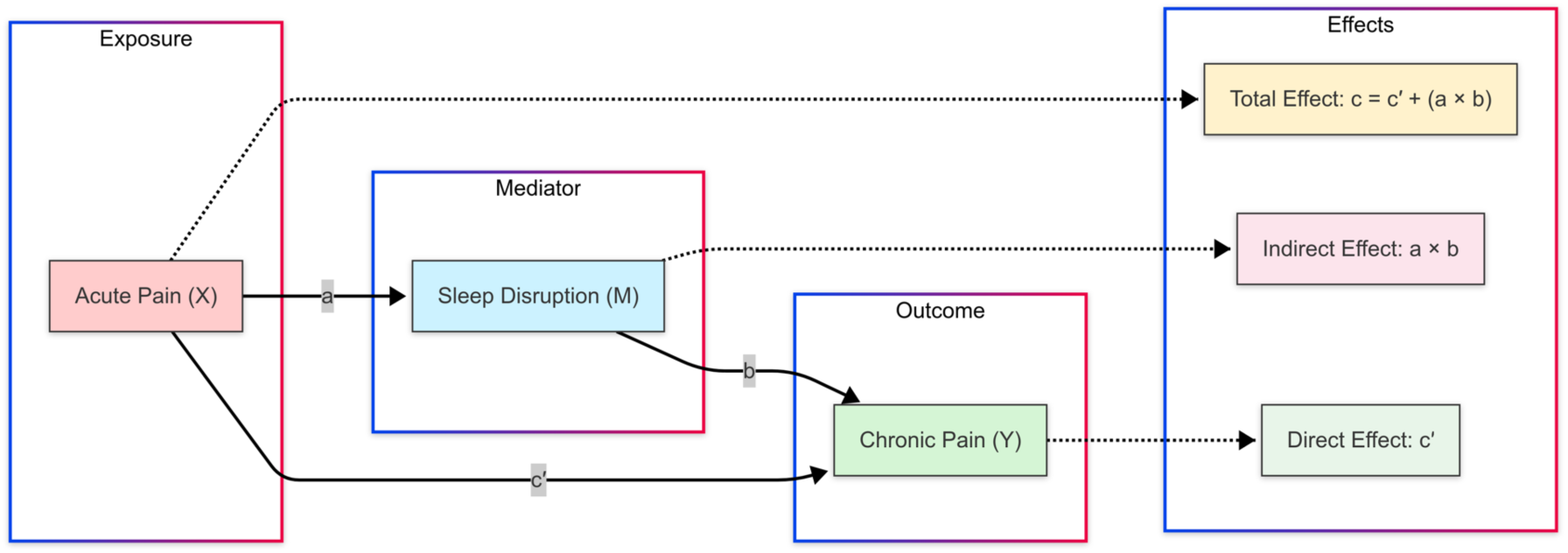
Proposed mediation model of early sleep disruption on the transition from acute to chronic postsurgical pain after caesarean delivery.

**Table 1.** Variables and association with chronic pain (all sites) after caesarean delivery; Univariate logistic regression; BPI-SF: Brief Pain Inventory Short Form used for pain outcomes; STAI= State Trait Anxiety Inventory; EPDS= Edinburgh Postnatal Depression Scale; P-values in **bold** are statistically significant. † Exploratory analysis using Firth’s penalised logistic regression.

| Variable | Odds Ratio (95% C.I.) | P-value |
| --- | --- | --- |
| Antenatal factors |  |  |
| Body Mass Index (BMI) | 1.14 (0.38 to 1.87) | 0.54 |
| Inpatient variables (24 to 48 h postoperative) |  |  |
| Least pain intensity over the past 24 h | 1.31 (1.09 to 1.60) | **0.006** |
| Average pain intensity over the past 24 h | 1.36 (1.08 to 1.76) | **0.012** |
| Worst pain intensity over the past 24 h | 1.25 (1.04 to 1.54) | **0.022** |
| Duration of time in severe pain in past 24 h | 1.21 (1.03 to 1.4) | **0.020** |
| Pain interference with sleep | 1.14 (1.01 to 1.31) | **0.044** |
| Pain interference with ability to feel in control | 1.13 (1.00 to 1.28) | 0.055 |
| Pain interference with general activities | 1.12 (0.98 to 1.27) | 0.088 |
| Number of acute pain sites ($\geq 2$ vs $\leq 1$) | 28.00 (0.0002 to 7.1) | 0.16† |
| Pain interference with ability to feed baby | 1.09 (0.96 to 1.23) | 0.179 |
| Pain interference with ability to hold baby | 1.09 (0.96 to 1.23) | 0.180 |
| 2 week variables |  |  |
| Average pain intensity | 1.43 (1.09 to 1.92) | **0.013** |
| Pain interference with general activities | 1.24 (1.03 to 1.51) | **0.028** |
| PROMIS Sleep Disturbance Short Form | 1.08 (1.01 to 1.15) | **0.029** |
| Pain interference with sleep | 1.20 (1.00 to 1.46) | 0.057 |
| Pain interference with ability to feed baby | 1.18 (0.98 to 1.44) | 0.087 |
| Pain interference with normal work | 1.14 (0.97 to 1.35) | 0.116 |
| Pain interference with ability to hold baby | 1.14 (0.95 to 1.38) | 0.152 |
| 3 month variables |  |  |
| EPDS | 1.09 (1.00 to 1.19) | **0.044** |
| PROMIS Sleep Disturbance Short Form | 1.05 (1.00 to 1.10) | **0.049** |
| STAI score | 1.04 (1.00 to 1.08) | **0.038** |

**Supplementary Table 1.** Sociodemographic and features of study population as per chronic pain group (n=100). Data are expressed as median [IQR] variable comparisons by pain group (pain present at 3 to 4 months Yes or No). P-values calculated using Mann–Whitney U test. * p < .05, ** p < .01, *** p < .001, . = trend (p < .10). Abbreviations: EPDS = Edinburgh Postnatal Depression Scale; STAI = State-Trait Anxiety Inventory; PROMIS = Patient-Reported Outcomes Measurement Information System. Using Pearson’s Chi-squared test with Yates’ continuity correction, there was no significant difference in the proportion of intrapartum versus planned caesarean deliveries between participants with and without chronic postsurgical pain (χ²(1) = 2.82, *p* = 0.093). For factor variables with small expected counts, Fisher’s Exact Test was used to compare CPSP vs no CPSP groups.

| Variable | Median [IQR] – No Chronic Pain | Median [IQR] – Yes Chronic Pain | P-value | Sig. |
| --- | --- | --- | --- | --- |
| **Baseline Characteristics** |  |  |  |  |
| Age | 33.5 [28, 37] | 34 [30, 37] | 0.624 |  |
| BMI | 30.9 [27, 34.8] | 28.3 [25.6, 33.2] | 0.176 |  |
| Race, n (%) |  |  |  |  |
| Asian | 15 (25%) | 16 (40%) | 0.12 |  |
| Native Hawaiian | 0 (0%) | 1 (2.5%) |  |  |
| Black or African American | 1 (1.7%) | 1 (2.5%) |  |  |
| White | 21 (35%) | 11 (28%) |  |  |
| More than 1 race | 2 (3.3%) | 3 (7.5%) |  |  |
| Unknown/ Not Reported | 1 (1.7%) | 2 (5.0%) |  |  |
| Other | 20 (33%) | 6 (15%) |  |  |
| Ethnicity, n (%) |  |  |  |  |
| Hispanic or Latino | 20 (33%) | 7 (18%) | 0.14 |  |
| Not Hispanic or Latino | 39 (65%) | 31 (78%) |  |  |
| Unknown/ not reported | 1 (1.7%) | 2 (5.0%) |  |  |
| Language, n (%) |  |  |  |  |
| English | 45 (75%) | 34 (85%) | 0.2 |  |
| Spanish | 15 (25%) | 6 (15%) |  |  |
| Education level, n (%) |  |  |  |  |
| Below Bachelor's degree | 17 (68) | 8 (32) | 0.404 |  |
| Bachelor's degree and above | 39 (55.7) | 31 (44.3) | 0.404 |  |
| Household income |  |  |  |  |
| \$1 – \$25,001 | 5 (8.3%) | 1 (2.5%) | 0.6 | |
| \$25,001 - \$50,000 | 6 (10%) | 2 (5.0%) | | |
| \$50,001 - \$100,000 | 4 (6.7%) | 2 (5.0%) | | |
| \$100,001 - \$150,000 | 5 (8.3%) | 2 (5.0%) | | |
| ≥ \$150,000 | 27 (45%) | 20 (50%) | | |
| Prefer not to answer | 13 (22%) | 13 (33%) |  |  |
| Insurance status, n (%) |  |  |  |  |
| Private | 39 (66%) | 31 (78%) | 0.2 |  |
| MediCal | 19 (32%) | 7 (18%) |  |  |
| Other | 1 (1.7%) | 2 (5.0%) |  |  |
| Relationship status |  |  |  |  |
| Single | 8 (13.3%) | 3 (7.5%) | 0.308 |  |
| Married | 43 (71.7%) | 33 (82.5%) |  |  |
| In a relationship | 9 (15%) | 3 (7.5%) |  |  |
| Prefer not to answer | 0 (0%) | 1 (2.5%) |  |  |
| Medical history, n (%) |  |  |  |  |
| Pre-existing pain condition | 3 (5%) | 2 (5%) | 1.0 |  |
| Chronic hypertension | 4 (6.7%) | 1 (2.5%) | 0.6 |  |
| Diabetes mellitus | 3 (5.0%) | 1 (2.5%) | 0.6 |  |
| Neurological | 3 (5.0%) | 2 (5.0%) | >0.9 |  |
| Psychiatric | 9 (15%) | 4 (10%) | 0.5 |  |
| Autoimmune | 3 (5.0%) | 0 (0%) | 0.3 |  |
| Renal | 1 (1.7%) | 0 (0%) | >0.9 |  |
| Haematological | 1 (1.7%) | 1 (2.5%) | >0.9 |  |
| Infectious | 0 (0%) | 1 (2.5%) | 0.4 |  |
| None | 33 (55%) | 26 (65%) | 0.3 |  |
| Other | 19 (32%) | 7 (18%) | 0.11 |  |
| Obstetric complication, n (%) |  |  |  |  |
| Preterm labor | 0 (0%) | 1 (2.5%) | 0.4 |  |
| Fetal anomaly | 1 (1.7%) | 0 (0%) | >0.9 |  |
| IUGR | 0 (0%) | 0 (0%) | >0.9 |  |
| LGA | 1 (1.7%) | 1 (2.5%) | >0.9 |  |
| Pre ET | 3 (5.0%) | 3 (7.5%) | 0.7 |  |
| Eclampsia | 0 (0%) | 0 (0%) | >0.9 |  |
| Gestational hypertension | 5 (8.3%) | 1 (2.5%) | 0.4 |  |
| Placenta accreta spectrum | 2 (3.3%) | 1 (2.5%) | >0.9 |  |
| PPROM | 0 (0%) | 1 (2.5%) | 0.4 |  |
| None | 14 (23%) | 7 (18%) | 0.5 |  |
| Other | 40 (67%) | 31 (78%) | 0.2 |  |
| Planned Caesarean Delivery, n (%) | 37 (68.5) | 17 (31.5) | - |  |
| Intrapartum Caesarean Delivery, n (%) | 23 (50) | 23 (50) | - |  |
| Gravida | 39 [38, 39] | 39 [38, 39] | 0.769 |  |
| Parity | 0 [0, 1] | 0.5 [0, 1] | 0.778 |  |
| **Adverse Life Events** |  |  |  |  |
| Adverse Childhood Experiences (ACEQ) | 0 [0, 1] | 0 [0, 2] | 0.741 |  |
| **Pain Catastrophizing** |  |  |  |  |
| Pain Catastrophizing Scale (PCS) | 5 [0.8, 16.5] | 5 [1, 12] | 0.862 |  |
| **Acute Pain (24 to 48 hours)** |  |  |  |  |
| Worst Acute Pain (NRS 0-10) | 6 [4, 8] | 7 [6, 8] | 0.025 | * |
| Duration of Time in Severe Pain (%) | 20 [0, 40] | 30 [20, 50] | 0.011 | * |
| Number of Acute Pain Sites | 4 [2, 7] | 4 [2, 6] | 0.362 |  |
| **Chronic Pain (3 to 4 months)** |  |  |  |  |
| Worst Pain (NRS 0-10) | 0 [0, 1] | 5 [4, 7] | <0.001 | *** |
| Number of Chronic Pain Sites | 1 [1, 1] | 2 [2, 4] | <0.001 | *** |
| **Pain Interference** |  |  |  |  |
| Sleep (24 to 48 hours) | 4 [1, 7] | 5 [3, 8] | 0.041 | * |
| Feeling in Control (24 to 48 hours) | 3.5 [0.8, 5.2] | 5 [1.5, 8.5] | 0.065 | . |
| **Sleep Disturbance (PROMIS)** |  |  |  |  |
| 2 weeks | 13.5 [0, 22] | 19 [11, 24.5] | 0.042 | * |
| 3 to 4 months | 16 [9.8, 22] | 19 [12, 25.5] | 0.110 |  |
| **Anxiety (STAI)** |  |  |  |  |
| 24 to 48 hours | 44 [41, 46] | 44 [43, 46] | 0.235 |  |
| 2 weeks | 46.5 [0, 49] | 47 [42.2, 49] | 0.378 |  |
| 3 to 4 months | 47 [46, 49] | 46.5 [44, 49] | 0.053 | . |
| **Depression (EPDS)** |  |  |  |  |
| EPDS (24 to 48 hours) | 4 [1, 8] | 3 [0, 9] | 0.534 |  |
| EPDS (2 weeks) | 2 [0, 7] | 0 [0, 8.2] | 0.738 |  |
| EPDS (3 to 4 months) | 1 [0, 5.2] | 3.5 [0, 8] | 0.045 | * |

**Supplementary Table 2.** Distribution of the number of pain sites among participants at three months postpartum. Pain sites were assessed across 45 predefined anatomical locations. The majority of participants reported pain affecting two or more sites.

| Pain Site Category | n | Percentage (%) |
| --- | --- | --- |
| 1 site | 2 | 4.3% |
| 2 sites | 24 | 52.2% |
| 3 or more sites | 20 | 43.5% |

**Supplementary Table 3.**
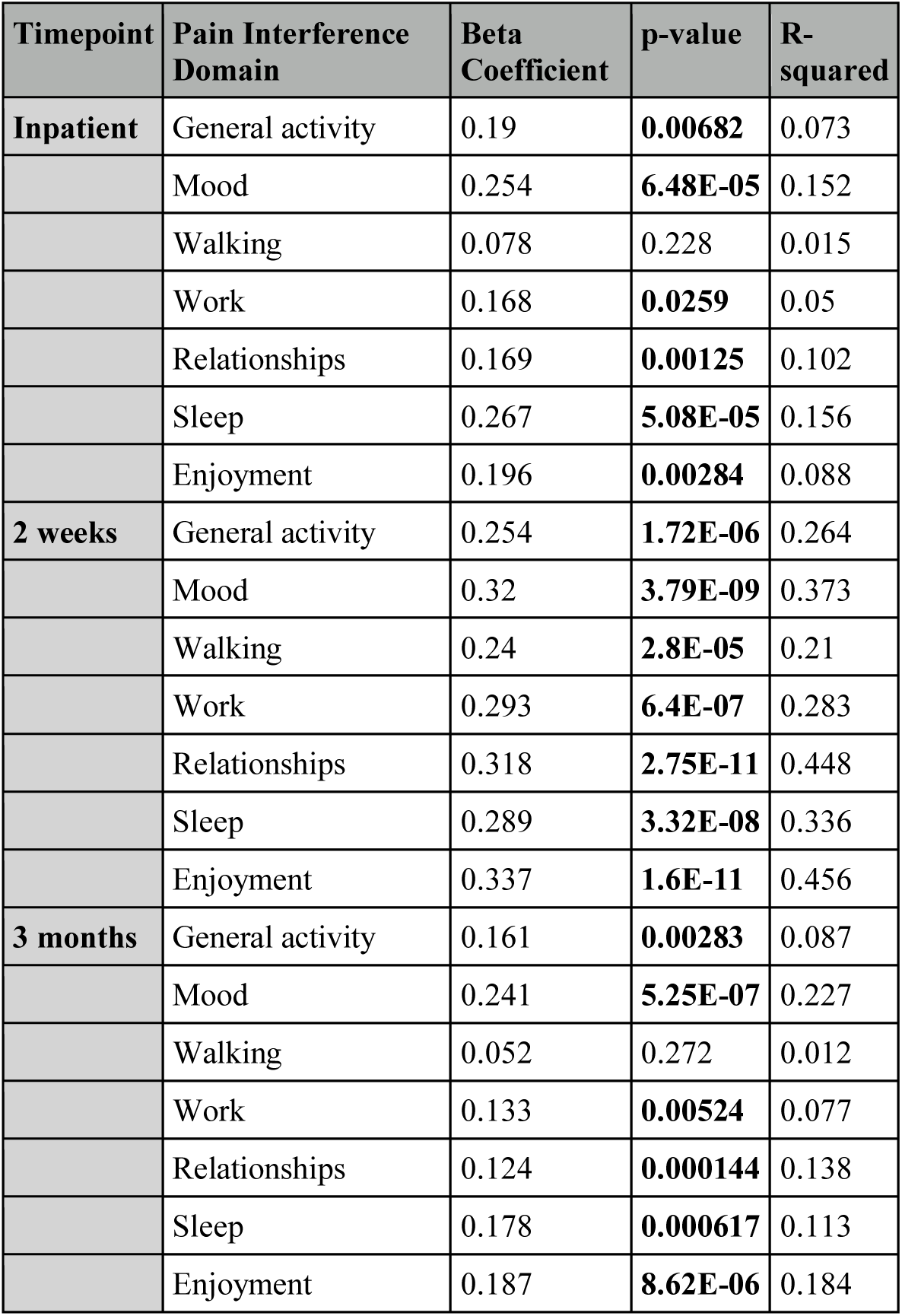
Associations Between Edinburgh Postnatal Depression Scale (EPDS) Scores and Pain Interference Domains at Inpatient, 2-Week, and 3-Month Timepoints. This table presents results from linear regression analyses evaluating the association between EPDS scores and patient-reported pain interference across seven functional domains: general activity, mood, walking ability, work, relationships, sleep, and enjoyment of life. Analyses were conducted at three postpartum timepoints: during inpatient stay, at 2 weeks, and at 3 months following caesarean delivery. For each model, the beta coefficient, p-value, and R-squared value are reported. Higher EPDS scores were consistently associated with greater pain interference, particularly in mood, sleep, relationships, and enjoyment.

**Supplementary Table 4.** Association between early pain catastrophising and psychological outcomes. Linear regression models examining the association between inpatient pain catastrophizing scores and anxiety, depression, and sleep disturbance outcomes at inpatient recovery, two weeks, and three months postpartum.

| Timepoint | Outcome | Beta (B) | p-value | R<sup>2</sup> |
| --- | --- | --- | --- | --- |
| Inpatient | Anxiety | 0.38 | **0.0005** | 0.078 |
| Inpatient | Depression | 0.29 | **<0.0001** | 0.303 |
| 2 weeks | Anxiety | 0.26 | 0.215 | 0.010 |
| 2 weeks | Depression | 0.18 | **<0.001** | 0.111 |
| 2 weeks | Sleep disturbance | 0.23 | **0.022** | 0.035 |
| 3 months | Anxiety | 0.38 | 0.067 | 0.022 |
| 3 months | Depression | 0.004 | 0.926 | 0.00006 |
| 3 months | Sleep disturbance | 0.18 | 0.10 | 0.018 |

**Supplementary Table 5.** AUC Values for Regularised Linear Models SPACE-Postpartum v0.9r.

| Method | AUC |
| --- | --- |
| Ridge (Python) | 0.759 |
| LASSO | 0.760 |
| Elastic Net | 0.760 |

**Supplementary Table 6.** Comparison of SPACE-Postpartum model variants for predicting chronic postsurgical pain at 3 months postpartum. Summary of model performance across four versions of the SPACE-Postpartum predictive model. The prototype binary model (v0.8) applied clinical cut-offs to each symptom domain. The primary model (v0.9r) used regularised linear terms and demonstrated good discrimination (AUC = 0.76). A quadratic ridge regression model (v0.9qr), achieved the highest apparent discrimination (AUC = 0.87) but may be overfitted given the sample size.

| Model Type | Description | AUC | Notes |
| --- | --- | --- | --- |
| v0.8 | Binary cut-off prototype | 0.64 | Simple, interpretable, low accuracy |
| v0.9 | Unregularised quadratic model | 0.83 | Unregularised model |
| v0.9r | Linear ridge (no quadratic terms) | 0.76 | Penalised, used for pilot tool |
| v0.9qr | Quadratic ridge regularised model | 0.87 | Best AUC, risk of overfitting |

**Supplementary Table 7.** Exploratory mediation analysis of early central nervous system-driven symptoms mediating the relationship between acute pain and chronic postsurgical pain after caesarean delivery. Mediation models examined whether 2-week sleep disturbance alone, or a composite SPACE symptom burden score (sleep disturbance + perceived loss of control), mediated the association between inpatient worst pain intensity (exposure) and 3-month worst pain intensity (outcome). ACME = average causal mediation effect (indirect effect); ADE = average direct effect; Total Effect = sum of ACME and ADE. All models were adjusted for age, body mass index (BMI), and parity. Estimates and 95% confidence intervals were generated via nonparametric bootstrapping with 1,000 simulations. Proportion mediated is presented as the ratio of ACME to Total Effect.

| Mediation Model | ACME (Indirect Effect) | 95% CI for ACME | ACME p-value | ADE (Direct Effect) | 95% CI for ADE | ADE p-value | Total Effect | 95% CI for Total Effect | Total Effect p-value | Proportion Mediated | 95% CI for Proportion Mediated | Proportion Mediated p-value |
| --- | --- | --- | --- | --- | --- | --- | --- | --- | --- | --- | --- | --- |
| **Sleep disturbance alone** | 0.00659 | -0.06097 to 0.070 | 0.824 | 0.35674 | 0.13300 to 0.60 | 0.002 | 0.36333 | 0.15734 to 0.60 | <0.001 | 0.01814 | -0.17104 to 0.26 | 0.824 |
| **SPACE score (sleep + control)** | 0.0304 | -0.0536 to 0.13 | 0.41 | 0.3329 | 0.1297 to 0.60 | <0.001 | 0.3633 | 0.1777 to 0.62 | <0.001 | 0.0837 | -0.1653 to 0.40 | 0.41 |

**Supplementary Table 8.** Published chronic postsurgical pain (CPSP) prediction models compared with SPACE-Postpartum. CV= cross-validation.

| Study | Surgery | Sample Size / Events | Predictors | Model Type | Penalisation | Performance (AUC) | Validation |
| --- | --- | --- | --- | --- | --- | --- | --- |
| Meretoja et al, 2017<sup>1</sup> | Breast cancer surgery | Dev: 860; Val: 453 & 231 (~14–20% CPSP) | Pre-op pain, BMI, axillary dissection, severe acute pain | Logistic regression risk score | No | External AUC ~0.74 | External ×2 |
| Dereu et al, 2018<sup>2</sup> | Breast cancer surgery | 200 (~30% CPSP 4mo) | Pre-op pain, depression, age <50, expected high pain | Logistic regression | No | Apparent AUC 0.81 | None (apparent only) |
| Papadomanolakis-Pakis et al, 2025<sup>3</sup> | Mixed elective surgeries | 960 (16% CPSP 3mo) | Demographics, surgical type, acute postop pain | Logistic regression (backward) | No (bootstrap shrinkage) | Corrected AUC ~0.75 | Internal bootstrap |
| van Driel et al 2022<sup>4</sup> | Mixed surgeries | Dev: 344; Val: 150 (22–29% CPSP) | Opioid use, bone surgery, pain POD14, cold sensitivity | Logistic regression | No | Dev AUC 0.82; similar external | External validation |
| Qian & Wang, 2024<sup>5</sup> | Total knee arthroplasty | 239 (23% CPSP) | Female sex, acute pain, knee function | Logistic regression nomogram | No | AUC 0.90 (internal only) | Apparent/internal |
| Liu et al, 2025<sup>6</sup> | Postpartum | 1,398 (27% CPSP 6mo) | Pain at 3d, BMI, parity, newborn weight, prenatal back pain | ML (LASSO + XGBoost) | Yes | AUC 0.88 | Internal CV |
| SPACE-Postpartum | Caesarean delivery | 100 (40% CPSP 3mo) | 5 early postpartum symptom domains (SPACE) | Ridge logistic regression | Yes | AUC 0.76 (95% CI 0.67–0.85), optimism-corrected 0.755 | Internal bootstrap + CV |

